# Artificial intelligence enables comprehensive genome interpretation and nomination of candidate diagnoses for rare genetic diseases

**DOI:** 10.1101/2021.02.09.21251456

**Authors:** Francisco M. De La Vega, Shimul Chowdhury, Barry Moore, Erwin Frise, Jeanette McCarthy, Edgar Javier Hernandez, Terrence Wong, Kiely James, Lucia Guidugli, Pankaj B Agrawal, Casie A Genetti, Catherine A Brownstein, Alan H Beggs, Britt-Sabina Löscher, Andre Franke, Braden Boone, Shawn E. Levy, Katrin Õunap, Sander Pajusalu, Matt Huentelman, Keri Ramsey, Marcus Naymik, Vinodh Narayanan, Narayanan Veeraraghavan, Paul Billings, Martin G. Reese, Mark Yandell, Stephen F. Kingsmore

## Abstract

**Background:** Clinical interpretation of genetic variants in the context of the patient’s phenotype is becoming the largest component of cost and time expenditure for genome-based diagnosis of rare genetic diseases. Artificial intelligence (AI) holds promise to greatly simplify and speed interpretation by comprehensively evaluating genetic variants for pathogenicity in the context of the growing knowledge of genetic disease. We assess the diagnostic performance of GEM, a new, AI-based, clinical decision support tool, compared with expert manual interpretation.

**Methods:** We benchmarked GEM in a retrospective cohort of 119 probands, mostly NICU infants, diagnosed with rare genetic diseases, who received whole genome sequencing (WGS) at Rady Children’s Hospital. We also performed a replication study in a separate cohort of 60 cases diagnosed at five additional academic medical centers. For comparison, we also analyzed these cases with commonly used variant prioritization tools (Phevor, Exomiser, and VAAST). Included in the comparisons were WGS and whole exome sequencing (WES) as trios, duos, and singletons. Variants underpinning diagnoses spanned diverse modes of inheritance and types, including structural variants (SVs). Patient phenotypes were extracted either manually or by automated clinical natural language processing (CNLP) from clinical notes. Finally, 14 previously unsolved cases were re-analyzed.

**Results:** GEM ranked >90% of causal genes among the top or second candidate, using manually curated or CNLP derived phenotypes, and prioritized a median of 3 genes for review per case. Ranking of trios and duos was unchanged when analyzed as singletons. In 17 of 20 cases with diagnostic SVs, GEM identified the causal SVs as the top or second candidate irrespective of whether SV calls where provided or inferred *ab initio* by GEM when absent. Analysis of 14 previously unsolved cases provided novel findings in one, candidates ultimately not advanced in 3, and no new findings in 10, demonstrating the utility of GEM for reanalysis.

**Conclusions:** GEM enables automated diagnostic interpretation of WES and WGS for all types of variants, including SVs, nominating a very short list of candidate genes and disorders for final review and reporting. In combination with deep phenotyping by CNLP, GEM enables substantial automation of genetic disease diagnosis, potentially decreasing the cost and speeding case review.

## Background

A central tenet of genomic medicine is that outcomes are improved when symptom-based diagnoses and treatments are augmented with genetic diagnoses and genotype-differentiated treatments. Worldwide, an estimated 7 million infants are born with serious genetic disorders every year [1]. The last decade witnessed a huge increase in the catalog of genes associated with Mendelian conditions, from about 2,300 in 2010 [2], to over 6,700 by the end of 2020 [3]. The translation of that knowledge, in conjunction with major improvements in WES and WGS and downstream analytical pipelines, has enabled increased rates of diagnosis, from about 10%, with single gene tests, to over 50% [4]. While limitations of read-alignment and variant calling were major obstacles to early clinical implementations of WES and WGS [5], they have been largely removed by algorithmic advances, hardware acceleration, and parallelization through cloud computing [6,7]. However, clinical interpretation of genetic variants in the context of the patient’s phenotype remains largely manual and extremely labor-intensive, requiring highly trained expert input. This remains a major barrier to widespread adoption, and contributes to continued, low rates of genomic testing for patients with suspected genetic disorders despite strong evidence for diagnostic and clinical utility and cost effectiveness [8].

The major challenge for genome-based diagnosis of rare genetic disease is to identify a putative disease-causing variant amid approximately four million benign variants in each genome, a problem akin to finding a needle in a haystack [9]. Clinical genome interpretation is, by necessity, performed by highly trained, scarce, genome analysts, genetic counselors and laboratory directors [10]. For an average of 100 variants for review per case [11] this translates to 50-100 hours of expert review per patient [10]. In practice, this has led to review of only about 10 variants per case, which somewhat defeats the purpose of genome-wide sequencing.

The genome interpretation process consists of iterative variant filtering, coupled with evidence-based review of candidate disease-causing variants [12]. This process was almost entirely manual until the advent of variant prioritization algorithms, such as Annovar [13] and VAAST [14], and was later improved by the integration of patient phenotypes in analyzes, e.g. Phevor [15], Exomiser [16], Phen-Gen [17], Phenolyzer [18], and more recently Amelie [19]. While these tools accelerate review times, their stand-alone performance has been insufficient for widespread clinical adoption, in part due to their inability to appropriately interpret structural variants (SVs). SVs account for over 10% of Mendelian disease [20,21], and about 20% of diagnoses in routine neonatal intensive care unit (NICU) [22] and pediatric patients [23]. Unified methods for prioritization of SVs, SNVs, and small indels is a fundamental requirement for further automation of genome interpretation.

The use of artificial intelligence (AI) has made significant inroads in healthcare [24] and a new class of genome interpretation methods [19,25–28] are being developed with the promise of removing the interpretation bottleneck for rare genetic disease diagnosis through electronic clinical decision support systems (eCDSS) [29]. Speed and accuracy of interpretation are particularly important for seriously ill children in the NICU [27], where diagnosis in the first 24-48 hours of life has been shown to maximally improve health outcomes [30]. The settings and extent to which AI facilitates diagnosis are still being investigated [27,28]. Issues include what types of AI methods are most suitable (e.g., Bayesian networks, decision trees, neural nets, etc. [31]); how they compare with current variant prioritization approaches in terms of accuracy; their diagnostic performance across different clinical scenarios and variant types; their potential to offer new forms of decision support; and how well they integrate with automated patient phenotyping and clinical decision making [27,28,32].

Algorithmic benchmarking in this domain is no simple matter. Hitherto, most attempts have used simulated cases (created by adding known disease-causing variants to reference exomes and genomes), included only a few cases, derived from a single center, or were limited to certain variant types [17,33,34]. Such benchmarking is inherently limited, as it is not representative of the true diversity of genetic diseases and variant types (e.g., by omitting cases with causal SVs), and provide no means to evaluate the impact of different sequencing and variant calling pipelines on performance.

Here we describe and benchmark the diagnostic performance of GEM, a new AI-based eCDSS, and compare it to current variant prioritization approaches in a diverse cohort of retrospective pediatric cases from the Rady Children’s Institute for Genomic Medicine (RCIGM). These cases are largely comprised of seriously ill NICU infants; all were diagnosed with Mendelian conditions following WGS (or, in a few cases, WES), using a combination of filtering and variant prioritization approaches. These real-world cases encompass the breadth of phenotypes and disease-causing variants, including pathogenic SVs. We then sought to replicate the diagnostic performance of GEM in a second set of affected, diagnosed and undiagnosed children outside the NICU. They were collected from five different academic medical centers, mostly consisting of WES, to examine the generalizability of GEM’s diagnostic performance to other sequencing, variant calling pipelines, and clinical settings. Finally, we re-analyzed a set of previously negative RCIGM cases to evaluate the ability of GEM to identify new diagnoses without suggesting numerous false positives that would lead to time consuming case reviews. Our results show that rapid, accurate, and comprehensive WGS- and WES-based diagnosis is achievable through integration of new data modalities with algorithmic innovations made possible by AI.

## Methods

### Patient selection, phenotyping, and specimen sequencing

#### Rady Children’s Hospital

119 cases with primary findings and 14 negative cases, consisting of mostly NICU admits, were sequenced as part of the rapid-WGS sequencing program at the Rady Children’s Hospital Clinical Genome Center. All subjects had a symptomatic illness of unknown etiology in which a genetic disorder was suspected (**Supp. Table 1**). WGS (or in a few instances WES) was performed as previously described [35,36]. Briefly, PCR-free WGS was performed to an average of 40x coverage in the Illumina HiSeq 2000, HiSeq 4000 and NovaSeq 6000 sequencers. Alignment and sequence variant calling where performed using the Illumina DRAGEN software, and copy number variation was generated through an approach that integrates the tools Manta [37] and CNVnator [38]. Structural variants were then filtered for recurrent artifacts observed in previous non-affected cases and only included in the input VCF file if they overlap a known disease gene (OMIM). All variants reported as primary findings where validated orthogonally. In some older cases, SV calling was not performed; any causal SVs therein where identified by an orthogonal CGH microarray or manual inspection of alignments.

**Table 1.** Characteristics of case cohorts. Benchmark cohort, 119 cases total; Validation cohort, 60 cases total. Grand total: 179 cases.

| Mode of Inheritance | Assay Type |  | Variant Type |  | Proband Sex |  | Pedigree Type |  |  |
| --- | --- | --- | --- | --- | --- | --- | --- | --- | --- |
|  | WGS | WES | SNV/Indel | SV | Male | Female | Single | Duos | Trios |
| *Benchmark cohort* |  |  |  |  |  |  |  |  |  |
| Autosomal Dominant | 70 | 11 | 66 | 15 | 36 | 45 | 35 | 6 | 40 |
| Autosomal Recessive | 27 | - | 23 | 4 | 14 | 13 | 9 | 1 | 17 |
| X-linked Dominant | 6 | - | 5 | 1 | 1 | 5 | 2 | - | 4 |
| X-Linked Recessive | 5 | - | 5 | - | 5 | - | 2 | 1 | 2 |
| Sub-Totals: | 108 | 11 | 99 | 20 | 56 | 63 | 48 | 8 | 63 |
| *Validation cohort* |  |  |  |  |  |  |  |  |  |
| Autosomal Dominant | 3 | 34 | 37 | - | 10 | 27 | 15 | 2 | 20 |
| Autosomal Recessive | 1 | 14 | 15 | - | 5 | 10 | 9 | - | 6 |
| X-linked Dominant | 1 | 5 | 6 | - | 3 | 3 | 1 | 3 | 2 |
| X-Linked Recessive | - | 2 | 2 | - | 2 | - | 1 | - | 1 |
| Sub-Totals: | 5 | 55 | 60 | 0 | 20 | 40 | 26 | 5 | 29 |

#### Boston Children’s Hospital

11 cases (all single-probands) from the Beggs Lab, Congenital Myopathy Research Program laboratory, and Manton Center for Orphan Disease Research at Boston Children’s Hospital were included in the analysis. Libraries (TruSeq DNA v2 Sample Preparation kit; Illumina, San Diego, CA) and whole exome capture (EZ Exome 2.0, Roche) were performed according to manufacturer protocols from DNA extracted from blood samples. WES was carried out on an Illumina HiSeq 2000. Reads were aligned to the GRCh37/hg19 human genome assembly using an in-house assembler. Variants were called using Gene Analysis Toolkit (GATK) version 3.1 or higher (Broad Institute, Cambridge, MA) and where Sanger confirmed by the Boston Children’s Hospital IDDRC Molecular Genetics Core Facility.

#### Christian-Albrechts University of Kiel

12 cases (all single-probands) from the Institute of Clinical Molecular Biology (IKMB) were included in the analysis. Illumina’s Nextera/TruSeq whole-exome target capture method was applied. WES was carried out on the Illumina HiSeq/NovaSeq platforms. Reads were aligned to the GRCh37/hg19 human genome assembly using BWA-MEM version 0.7.17 and variants called using GATK version 4.1.6.0 (Broad Institute, Cambridge, MA).

#### HudsonAlpha Institute for Biotechnology

Three cases (two trios and a single-proband) from the Clinical Services Laboratory at HudsonAlpha Institute for Biotechnology were included in the analysis. WGS was carried out on an Illumina HiSeq X. Reads were aligned to the GRCh37/hg19 human genome assembly followed by variant calling using the Illumina DRAGEN software version 3.2.8 (Illumina, Inc. San Diego, CA).

#### Translational Genomics Research Institute

23 cases (including singletons, duos, trios and quads) from the Center for Rare Childhood Disorders at The Translational Genomics Research Institute (TGen) were included in the analysis. WES or WGS sequencing was carried out on an Illumina HiSeq 2000, HiSeq 2500, HiSeq 4000, or NovaSeq6000. For WES, the Agilent SureSelect Human All Exon V6 or CRE V2 target capture method was applied. Reads were aligned to reference GRCh37 version hs37d5 and variants called using GATK Haplotype caller version 3.3-0-g37228af (Broad Institute, Cambridge, MA).

#### Tartu University Hospital

11 cases from Tartu University Hospital in Estonia that had undergone Nextera Rapid Capture Exome Kit-i (Illumina Inc.) target capture method was applied. WES was carried out on an Illumina HiSeq2500 sequencer. Reads were aligned to the GRCh37/hg19 human genome assembly using BWA-MEM version 0.5.9 and variants called using GATK Haplotype caller version 3.4 (Broad Institute, Cambridge, MA).

### Variant annotation and data sources

All analyses where performed based on the GRCh37 human genome assembly. Variant consequences and annotations where obtained with VEP v.95 [39] utilizing ENSEMBL transcripts version 95 (excluding non-coding transcripts) and selecting the canonical transcript for analysis. Transcript specific prediction for evaluating variant deleteriousness were calculated with VVP [40], which were also used as input for VAAST [14]. Variants where annotated with ClinVar (20200419) [41] ensuring exact position and base match. Gene-conditions where extracted from OMIM (2020_07) [42] and HPO (obo file, 2020-08-11) [43]. Gene symbols where harmonized using ENSEMBL and HGNC databases controlling for synonymous gene symbols.

### AI-based Disease Gene and Condition Prioritization

AI-based prioritization and scoring of candidate disease genes and diagnostic conditions was performed using GEM in the commercially available Fabric Enterprise platform (Fabric Genomics, Oakland, CA). GEM inputs are genetic variant calls in VCF format and case metadata, including (optional) parental affection status, and patient (proband) phenotypes in the form of Human Phenotype Ontology (HPO) terms. The VCF files can include “small variants” (single nucleotide, multiple nucleotide, and small insertion/deletion variants), and (optionally) structural variants (insertion/deletions of over 50bp, inversions, and/or copy number variants with imprecise ends). This information can be provided via an application programming interface or manually in the user interface. Data analysis is typically carried out in minutes depending on inputs. GEM outputs are displayed in an interactive report (**Supp. Fig. 1**) that includes a list of candidate genes ranked by the GEM gene score (see below), detailed information of patient variants present in each candidate gene, and conditions associated with each candidate gene ranked by GEM’s condition match (CM) score (explained below).

**Figure 1.**
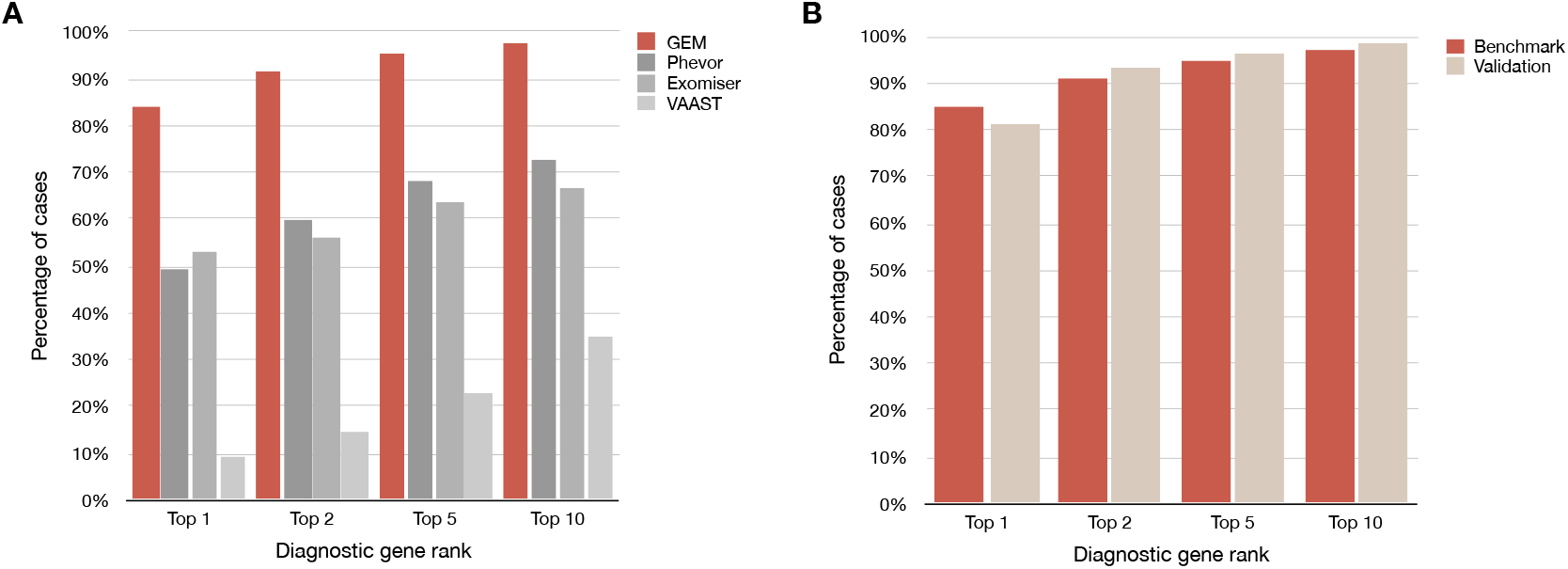
The diagnostic sensitivity of GEM was greater than the variant prioritization methods. Phevor, Exomiser and VAAST. (**A**) Proportion of the benchmark cohort of 119 cases where the true causal gene (or variant in the case of causal SVs) was identified among the top 1st, 2nd, 5th, or 10th gene candidates. Patient phenotypes were extracted manually from medical records by clinicians and provided as HPO term inputs to GEM, Exomiser and Phevor. VAAST only considers variant information. It should be noted that GEM and Phevor ranks correspond to genes, which may include one or two variants (the latter in the case of a compound heterozygote), whereas Exomiser and VAAST ranks were for single variants. In the case of compound heterozygotes, the rank of the top-ranking variant is shown for Exomiser and VAAST. (**B)** Comparison of GEM performance in the benchmarking cohort (excluding SV cases) versus the validation cohort (comprised of 60 rare genetic disease cases from multiple sources).

GEM aggregates inputs from multiple variant prioritization algorithms with genomic and clinical database annotations, using Bayesian means to score and prioritize potentially damaged genes and candidate diseases. Briefly, the algorithm parametrizes itself using the proband’s called variants as one-time, run-time training data, inferring the states of multiple variables directly from the input variant distribution, e.g., sex. Additional static training parameters were derived from the 1000 Genomes Project [44] and CEPH [45] genome datasets. GEM reevaluates genotype calls and quality scores considering read support, genomic location, proband sex and potentially overlapping SVs, augmenting the genotype calls with more nuanced posterior probabilities, computing ploidy for each variant. GEM also computes the likelihood that the proband belongs to any of several different ancestry groups using the input genotypes together with gnomAD sub-population variant frequency data [46]. The probabilities of other, internal, variables, conditioned on each state (sex and ancestry, etc.) are then obtained using naive Bayes, controlling for non-independence of variables by calculating a correlation matrix at run time using the proband’s data.

GEM’s gene scores are Bayes Factors (BF) [47]. Analogous to the likelihood ratio test, a Bayes Factor presents the log_10_ ratio between the posterior probabilities of two models, summarizing the relative support for the hypothesis (in this case) that the prioritized genotype damages the gene in which it resides and that explains the proband’s phenotype versus the contrapositional hypothesis that the variant neither damages the gene nor explains the proband’s phenotype. In keeping with established best practice [47], a log_10_ Bayes Factor between 0 and 0.69 is considered moderate support, between 0.69 and 1.0 substantial support, between 1.0 and 2.0 strong support, and above 2.0, decisive support. A score less than 0 indicates that the counter hypothesis is more likely. For calculating the Bayes posterior p(M|D), the probability of the data given a model (pD|M) is derived from GEM’s severity scoring protocol, which includes input from the VAAST and VVP algorithms, and any available prior variant classifications from the ClinVar database. This model is conditioned upon sex, ancestry, feasible inheritance model, gene location, and gene-phenotype priors derived by seeding the provided patient HPO terms to the HPO ontology graph and subsequently obtaining priors for all genes in the HPO and GO ontologies by belief propagation using Phevor’s previously described Bayesian network-based algorithm [15]. The prior probability for the model (pM) is based upon known disease associations in the Mendelian conditions databases OMIM and/or HPO with the gene in question.

GEM’s Bayes Factor-based scoring system is designed for ease of explanation and to speed interpretation. GEM scores are not intended to be definitive, rather they are designed to provide guidance for succinct case reviews carried out by clinical geneticists. Thus, GEM outputs also include several additional scores that provide additional guidance and improve explainability. GEM gene scores, for example, are accompanied by VAAST [14], VVP [40], and Phevor [15] posterior probabilities, conditioned upon the potentially confounding variables of proband sex, gene location and ancestry, together with common variant genomic and clinical annotations (**Supp. Fig. 1**). These scores further ease interpretation, as they allow users to assess the major drivers of a GEM score and their relative contributions to it.

GEM also provides means to assess the Mendelian conditions associated with putative disease-causing genes as possible diagnoses via its condition-match (CM) scores. Like gene scores, CM scores are Bayes Factors, and are derived from the log_10_ ratio of the posterior probability that HPO phenotype associations for a given Mendelian condition’s HPO are consistent with the proband’s phenotype versus the contrapositional hypothesis. For these calculations, the probability of the data, p(D|M), is determined using Phevor’s Bayesian network algorithm to obtain a probability for each disease, conditioned upon the proband’s phenotype. The prior probability for the model, p(M), is the probability that one or more genes associated with the Mendelian condition (as documented in OMIM and/or HPO) contain a damaging genotype as ascertained by GEM’s severity scoring protocol. Condition match scores are displayed alongside each gene-associated condition for review (**Supp. Fig 7**).

### Structural variant scoring and *ab initio* inference by GEM

At run time, GEM infers *ab initio* the existence of SVs, their coordinates, and their copy numbers (ploidy) in a probabilistic fashion using SNVs, sort indel calls, read depths, zygosity and gnomAD frequency data. GEM searches the proband’s genotypes for evidence of three types of SV: deletions, duplications and CNVs. Regions exhibiting loss of heterozygosity (LOH), for example, are used as evidence heterozygous deletions. Genomic spans lacking expected variants, the signature of homozygous deletions, are identified using gnomAD population frequencies [46] to derive point estates that a given gnomAD variant would or would not be ascertained given its population frequency. Further evidence for duplications and deletions is derived from read support, e.g. approximately integer increases or decreases in depth across a region provide support for copy number variation. Point estimates at each site of a small variant call are further conditioned upon provided variables, such as genotype qualities, and inferred ones, such as sex and ancestry, to obtain more refined estimates. High scoring segments and their maximum likelihood start and end coordinates are identified using a Markov model [48]. The results are used to determine the degree of support for external SV calls, and as the basis for GEM’s own SV calls. For ease of reporting, *ab initio* SV calls that overlap an external SV call (default minimum reciprocal overlap of 33%) are replaced in the output by the external SV call as long as they still overlap the relevant scored genes.

### Benchmarking Variant Prioritization with VAAST, Phevor, and Exomiser

We used the Snakemake software [49] to create a workflow that analyzes cases with the VAAST, Phevor, and Exomiser algorithms. The pipeline starts with a VCF file, family structure, affection status, and HPO terms, and produces a test file that includes the output for each of the algorithms. VVP scores where obtained as described above and provided to VAAST as input. VAAST was provided pedigree information and affection status and was run in both dominant and recessive modes with results aggregated. Gene ranks for VAAST are reported for the highest scoring occurrence of the gene from aggregated outputs. Phevor was provided with HPO terms and VAAST scores as inputs. Ranks where selected as described for VAAST.

Exomiser [16] benchmark analyses were run with the same configuration used in the 100,000 Genomes Project [50], specifically: 1) using the GRCh37 genome assembly; 2) analyzing autosomal and X-linked forms of dominant and recessive inheritance; 3) allele frequency sources from the 1,000 Genomes Project, TopMed, UK10K, ESP, ExAC, and gnomAD (except Ashkenazi Jewish); 4) pathogenicity sources from REVEL and MVP; and 5) including the steps failedVariantFilter, variantEffectFilter (remove non-coding variants), frequencyFilter with maxFrequency = 2.0, pathogenicityFilter with keepNonPathogenic = true, inheritanceFilter, omimPrioritiser and hiPhivePrioritiser.

Exomiser was considered to have identified the diagnosed gene when it was scored as a candidate for any of the utilized modes of inheritance. None of the tools in this analysis were provided a target mode of inheritance (as it is unknown), and so the diagnostic gene rank for Exomiser was determined from its rank within the combined gene candidate list from all modes of inheritance (i.e., the same procedure used for VAAST and PHEVOR). The ranks within the combined list of candidate genes were generated by sorting gene-level candidates from all modes of inheritance on the Exomiser combinedScore in descending order with each candidate-gene only added to the list on its first, highest scoring occurrence. Exomiser variant level scoring was not considered for determining candidates or ranking. All Exomiser analyses on the benchmark dataset ran to completion and successfully produced output; however, in 18 cases, Exomiser did not identify the true positive diagnostic gene as a scored candidate (i.e., it was absent from its output). A similar phenomenon was observed in 4 cases using VAAST. For both tools, these cases were considered false negatives.

### Impact of deep phenotypes derived from clinical NLP

The utility of HPO terms was investigated by rerunning all analyses from the benchmark cohort with three sets of HPO terms. For each case an HPO terms list was provided that included HPO terms manually curated by the analysis team when the case was originally solved. A second set of HPO terms was generated from NLP analysis of clinical notes related to the case using the CLiX ENRICH software (Clinithink, Alpharetta, GA) [28]. A randomized set of HPO terms was generated for each case whereby the number of HPO terms from the CliniThink analysis case was held constant, and alternate terms were randomly selected from the entire corpus of HPO terms across all samples with each selection probability determined by the number of times that term occurred in the corpus.

## Results and Discussion

### GEM AI outperforms variant prioritization approaches

We benchmarked GEM, an AI-based eCDSS, using a cohort of 119 pediatric retrospective cases from Rady Children’s Institute for Genomic Medicine (RCIGM; benchmark cohort). Most of these were critically ill NICU infants who received genomic sequencing for diagnosis of genetic diseases. All had been diagnosed with one or more Mendelian conditions using a combination of manual filtering and variant prioritization approaches (“Methods”). To further validate performance, we also analyzed a second cohort comprised of 60 non-NICU, rare disease patients from five different academic medical centers (validation cohort). Finally, we re-analyzed a set of 14 previously analyzed probands that had remained undiagnosed by WGS. Our goal was to evaluate the ability of GEM to identify new diagnoses in these previously negative cases, without providing false positive findings that would result in time consuming case reviews. To provide context for our performance benchmarks, we also ran three commonly used variant prioritization tools: VAAST [14], Phevor [15], and Exomiser [16].

The benchmark and validation cohorts included singleton probands, parent-offspring trios, different modes of inheritance, and both small causal variants (SNVs, and small insertions or deletions, indels; **Table 1; Supp. Table 1)** and large structural variants (SV), some of which were causative (**Table 2)**. In these retrospective analyses, we considered the variants, disease genes and conditions that were included as primary findings in the clinical report as the “gold standard” truth set.

**Table 2.**
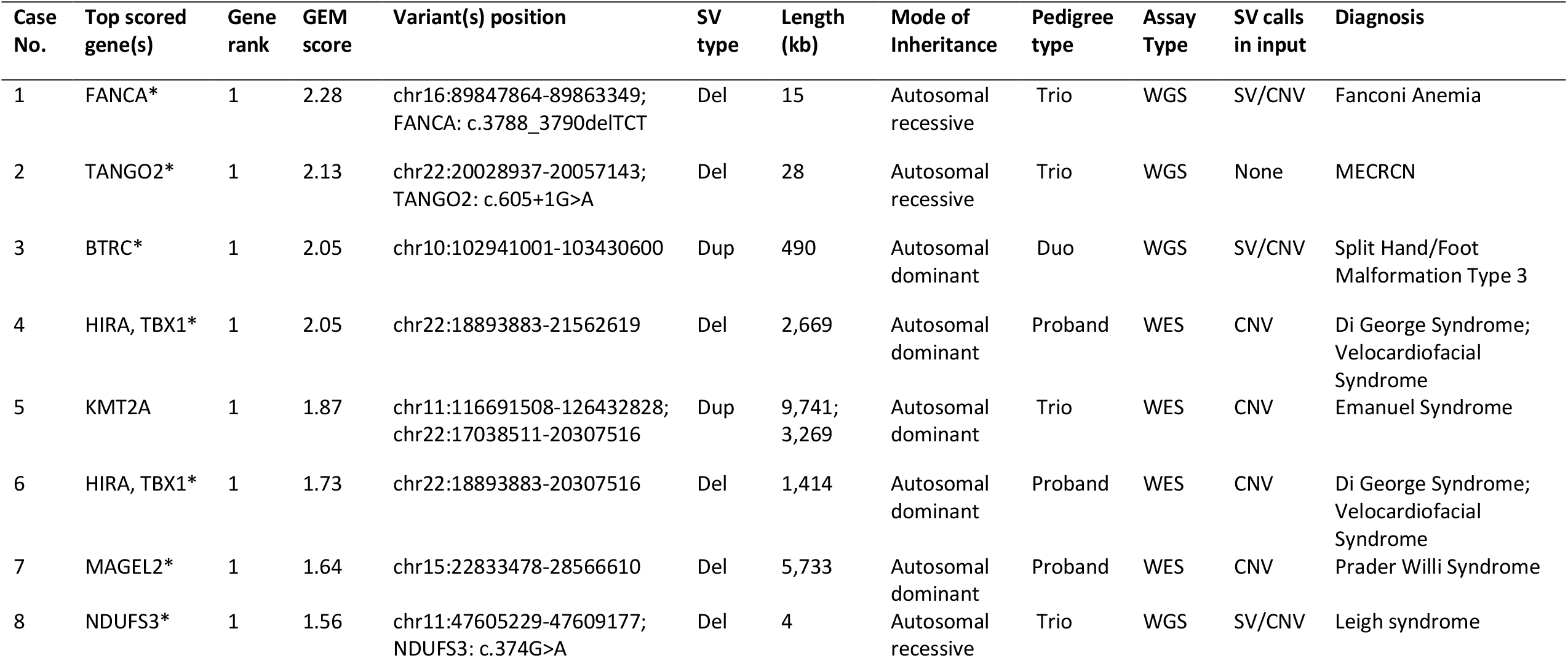
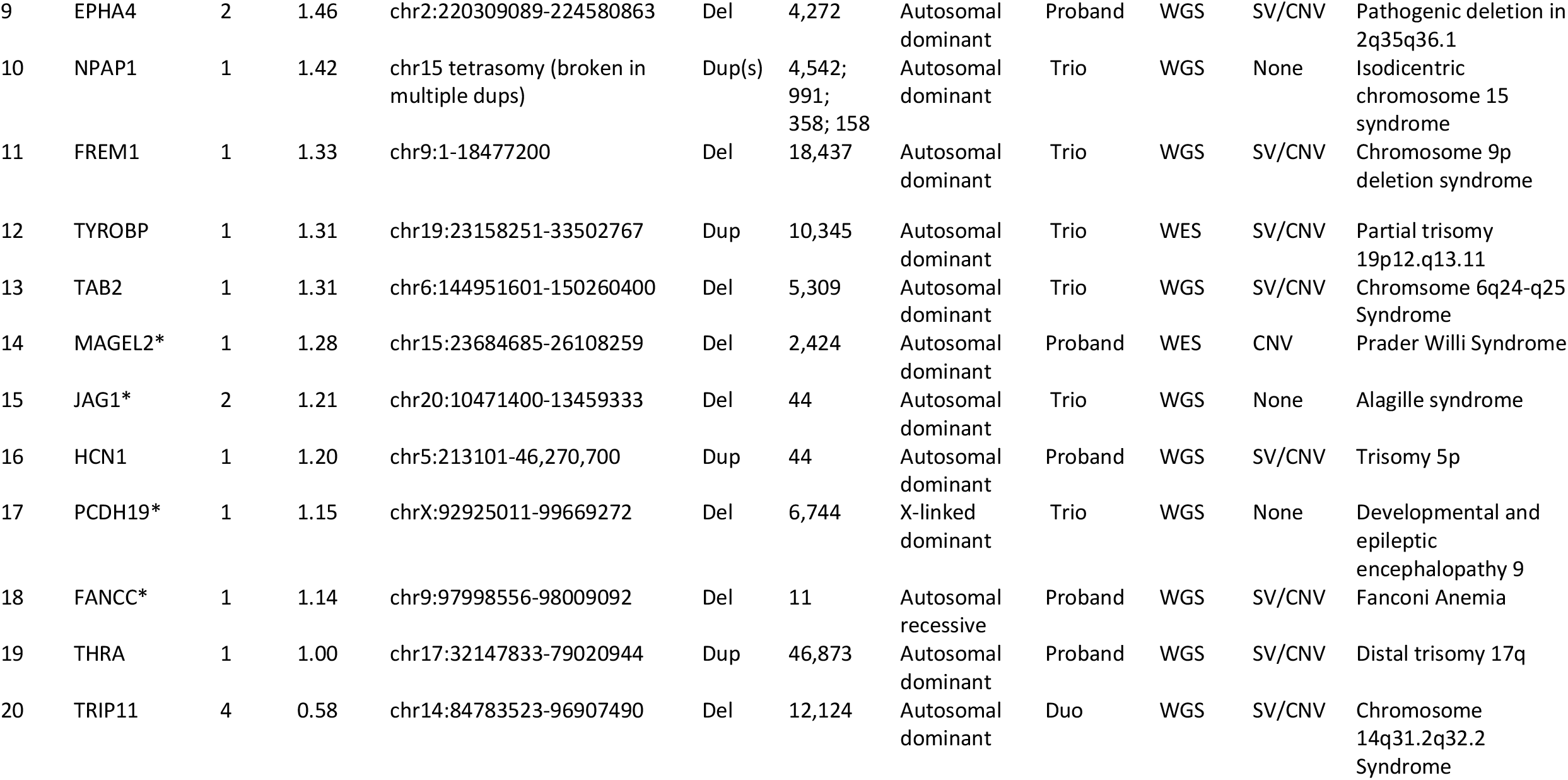
Diagnostic structural variants identified by GEM in the benchmarking cohort (20 out of 119 cases). Structural variants are ranked by GEM based on the genes harbored by the variant and presented alongside other ranked genes with coding SNVs or small indels based on the top scored gene. The asterisk indicates genes that in the literature are candidates for the phenotype of the diagnostic disease/syndrome (as described in OMIM). The results show that GEM can analyze both deletions (del) and duplications (dup) of sizes as small as 4Kb and up to entire chromosome arms, diverse modes of inheritance, pedigree structure, and from either WGS or WES assay data. GEM also automatically identified compound heterozygotes between SVs and SNV/indels (cases 1, 2, and 8). Input SV calls can include breakpoint-based calls (here “SV”), or imprecise CNV calls based on read depth analysis. Notably, GEM can also infer SVs directly from the small variant data when external SV calls are not provided (cases 2, 10, 15, and 17), and score them appropriately, identifying diagnostic variants that in the original cases where found by microarrays and not by sequencing.

GEM gene scores are Bayes Factors (BF) [51]; these were used to rank gene candidates (**Supp. Fig. 1**). BFs are widely used in AI, as they concisely quantify the degree of support for a conclusion derived from diverse lines of evidence. In keeping with established practices [51], a BF of 0 – 0.69 was considered moderate support, 0.69 – 1.0 substantial support, 1.0 – 2.0 strong support, and above 2.0, decisive support [51]. Scores less than 0 indicated support for the counter hypothesis – that variants in that gene were not causal for the proband’s disease. GEM outputs also included several annotations and metrics that provided additional, supportive guidance for subsequent expert case review (**Supp. Fig. 1**). Experience has shown that such guidance is critical for adoption by experts who wish to review the evidence supporting automated variant assertions. These included VAAST, VVP and Phevor posterior probabilities, conditioned upon proband sex, gene location and ancestry. Annotations included variant consequence, ClinVar database pathogenicity assessments, and OMIM conditions associated with genes. This metadata enabled expert users to review the major contributions underpinning a final GEM score. Moreover, GEM prioritizes diplotypes, rather than variants, which speeds interpretation of compound heterozygous variants in recessive diseases (**Supp. Fig. 1B**). Comparison of the diagnostic performance of GEM to variant priortization methods utilized ranking of the correct diagnostic gene. We assumed that in the case of compound heterozygotes, variant prioritization methods such Exomiser, would rank one variant of the pair highly, leading to identification of the other upon manual review (“Methods”).

GEM ranked 97% of previously reported causal gene(s) and variant(s) among the top 10 candidates in the 119 benchmark cohort cases. In 92% of cases, it ranked the correct gene and variant in the top 2 (**Fig. 1A**). By comparison, the next best algorithm, Phevor, identified 73% of causal variants in the top 10 candidates and 59% in the top 2. GEM, Phevor and Exomiser prioritize results by patient phenotypes (provided as HPO terms) in addition to variant pathogenicity, whereas VAAST only utilizes genotype data, explaining its lower performance. Thus, these data also highlight that patient phenotypes improve the diagnostic performance of automated interpretation tools.

Next, we investigated whether the diagnostic performance of GEM extended to Mendelian diseases other than those of NICU infants, such as patients with later disease onset, less severe presentations, or with data produced by other variant calling pipelines or outpatient genetic clinics. For these analyses, we compiled a validation cohort largely consisting of WES cases from five different academic medical centers (**Table1; Supp. Table 2**). The diagnostic performance of GEM in the validation cohort was almost identical to that in the benchmark cohort (**Fig. 1B**). These data demonstrated that the diagnostic performance of GEM was not dependent of disease severity, age of onset, or genomic sequencing or variant detection methods.

An implication of these findings is that GEM achieved 97% recall (true positive rate) by review of 10 genes, whereas the other tools had ≤ 78% recall by similar review (**Fig. 1,Supp. Fig. 2**). In part, this difference reflected the unique ability of GEM to prioritize SVs. Excluding SV cases, GEM, Phevor and Exomiser achieved recall of 97%, 83% and 76%, respectively, by review of 10 genes (**Supp. Fig. 3A**). Furthermore, VAAST and Exomiser failed to provide rankings for 4 and 18 true positive variants, respectively. Exclusion of false negatives and SV cases increased the top 10 recall of Exomiser to 93% (**Supp. Fig. 3B**), in agreement with previous reports [52]. These data show the importance of including all types of cases and causal variants in benchmarking to avoid overestimation of diagnostic performance in real-world clinical applications.

**Figure 2.**
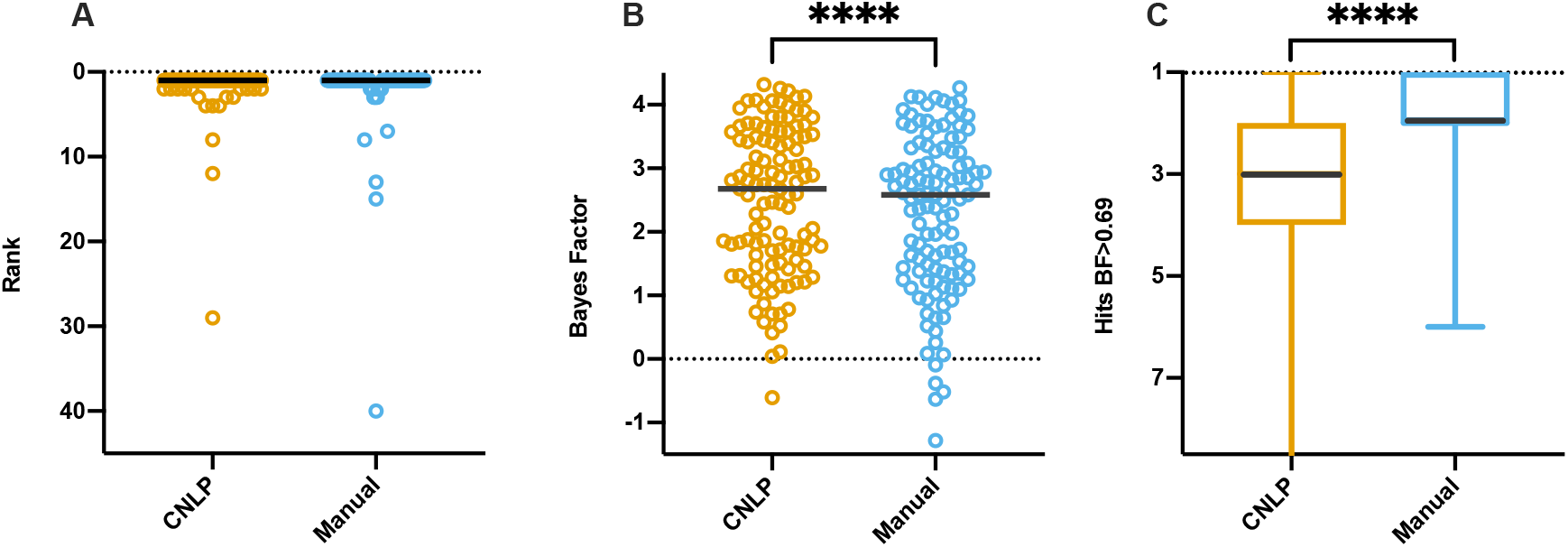
Comparison of GEM performance with manually curated and CNLP-derived HPO terms. Distribution of ranks for causal genes, (**A**); GEM Bayes Factors for causal genes, (**B**); and number of candidates (hits) at BF≥0.69 threshold (moderate support), (**C**). The black line in the graphs denotes the median. The asterisks represent statistical difference between the groups with p<0.0001 from a two-tailed Wilcoxson matched-pairs signed rank test (ranks showed no statistically significant difference).

**Figure 3.**
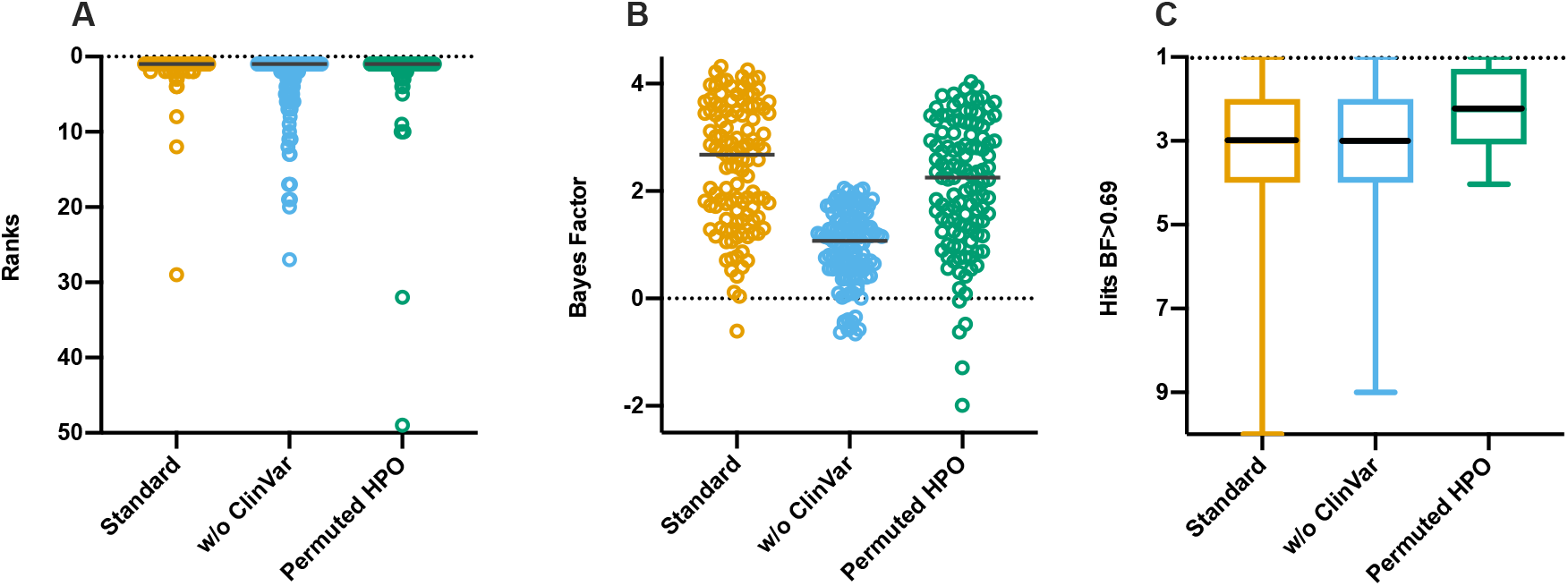
Impact of missing data and mis-phenotyping on GEM performance. Causal gene rank, (**A**); Bayes Factors for causal genes, (**B**); and number of candidates (hits) above gene BF≥0.69 threshold (moderate support), (**C**) under standard conditions, withdrawing ClinVar information, and permuting HPO terms extracted by CNLP. The black line in the graphs denotes the median.

### Scoring of structural variants increases diagnostic rate

Another major barrier to the incorporation of SV calls into genome diagnostic interpretation, whether manual or using eCDSS, is their low precision (high false positive, FP, rates) in short read alignments, with typical FP rates of 20-30% [53,54]. This leads to overwhelmingly time-consuming, manual assessment of event quality and significance for large numbers of SVs. GEM minimizes the effect of low precision by scoring SVs either with SV calls provided in the proband’s input VCF file, and/or by inferring *ab initio* their existence from metadata associated with SNV and indel calls (“Methods”; see below). The benchmark cohort included 20 cases in which SVs were reported to be causative, reflecting a similar incidence to that in real world experience (**Fig. 1A. Table 2)** [20–23]. In 17 of these, the causative SV was ranked first by GEM. In two, it was ranked second, and in one it was listed fourth, demonstrating that GEM retains adequate diagnostic performance with imprecise SV calls. The disease-causing SVs in the benchmark set ranged from small (4kb) to very large (e.g., entire chromosome arms). In three cases the diagnosis was of an autosomal recessive disorder in which the SV was compound heterozygous with a SNV/indel. In each, GEM integrated the two variants correctly, automatically identifying the causative diplotypes (**Supp. Fig. 5**). With regard to the diagnostic specificity of GEM, the mean and median number of gene-candidates for these probands with BF > 0 (any support) was 8.7 and 9.5, respectively, which was similar to probands whose VCF files contained no SVs, causative or otherwise.

**Figure 5.**
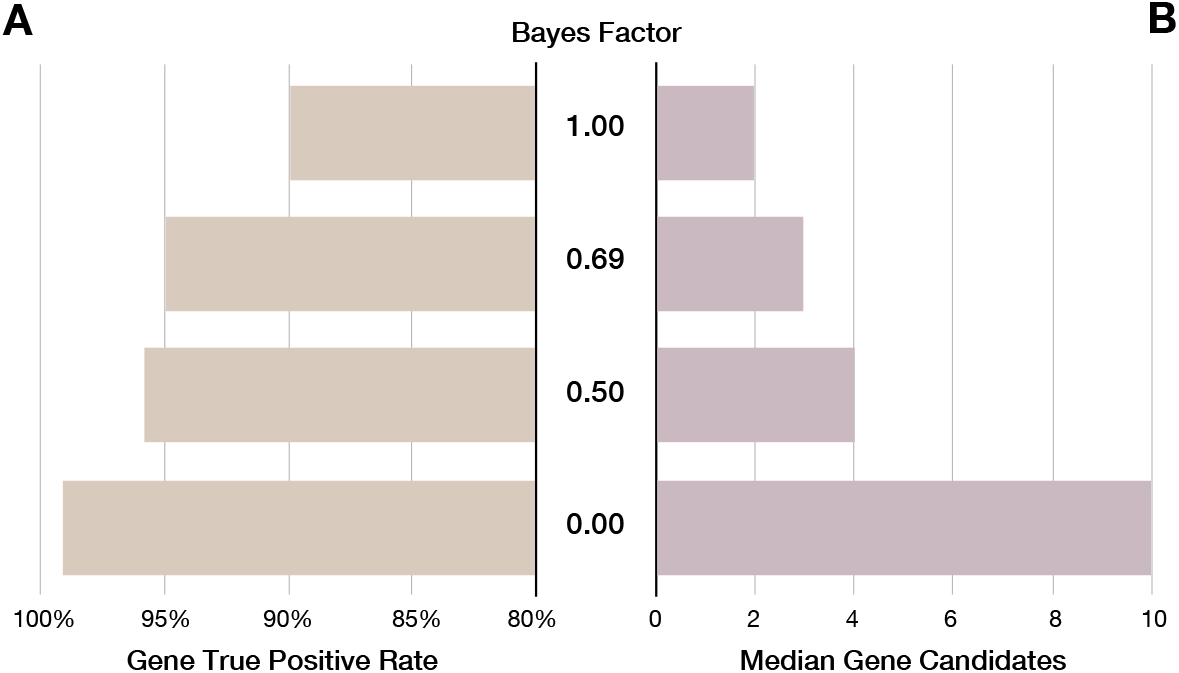
Trade-off between GEM gene scores, maximal true positive rates, and number of candidates for review. GEM gene scores are Bayes factors (BF) that can be used speed case review. **(A**) Gene maximal true positive rate achieved at the different BF thresholds (Y-axis). (**B**) Median number of candidate genes for review at each BF threshold. As the BF threshold is decreased, true positive rate increases while, the number of candidates to review remains manageable. Input HPO terms for this analysis where extracted by CNLP.

Large SVs frequently affect more than one gene. For consistency with other variant classes, genes within multigenic SVs are grouped and sorted by GEM based upon the gene-centric Bayes Factor score associated with the overlap of the proband phenotype and known Mendelian disorders (if any) associated with them (“Methods”). For example, **Supp. Fig. 4** shows an example that highlights the practical utility of prioritizing genes harboring causative SVs together with SNVs and short indels in the same report, rather than separately cross-referencing with databases of microdeletion syndromes [55]. While it is often unknown which genes harbored in a pathogenic SV are causal for microdeletion/microduplication syndromes, GEM’s gene-by-gene rankings typically agreed with causal gene candidates suggested by the literature (asterisks in **Table 2**).

**Figure 4.**
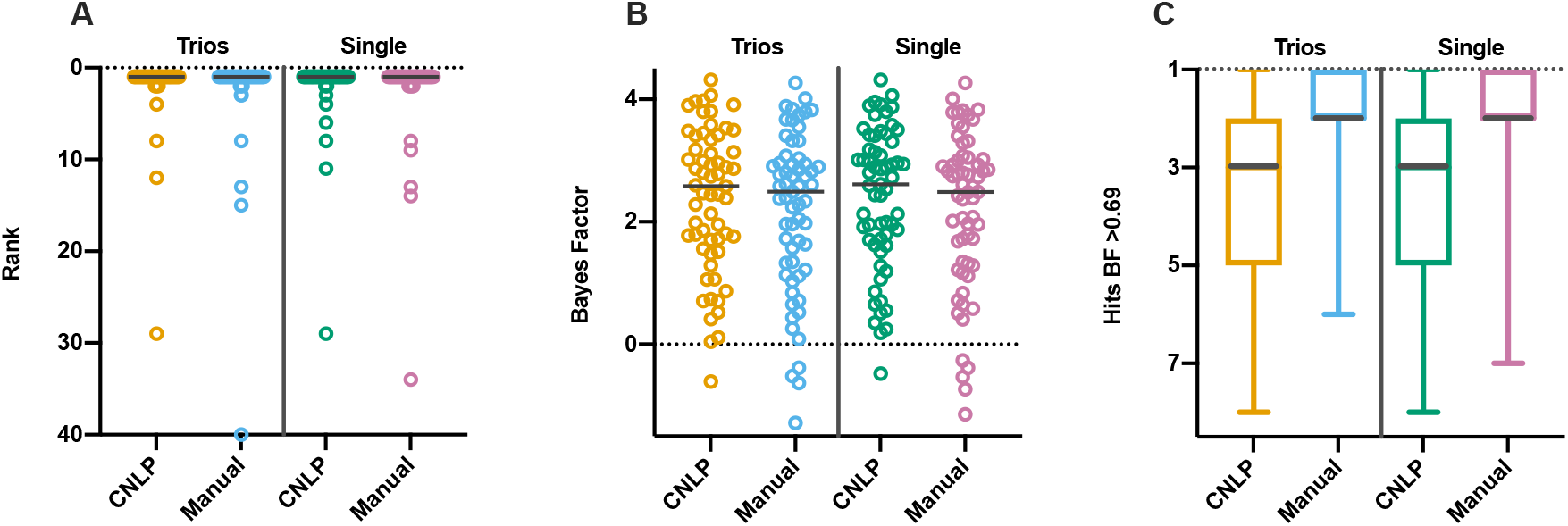
Comparative performance of parent offspring-trios or duos versus singleton probands. Causal gene rank, (**A**); Bayes Factors, (**B**); and number of candidates (hits) above gene BF ≥ 0.69 moderate support), (**C**) for 63 cases analyzed as parent-offspring trios (n=59) or duos (n=4), as compared with analysis as single probands, using both manually curated or CNLP-derived HPO terms. The black line in the graphs denotes the median. No statistically significance difference between the any Manual/CNLP groups was found between trios versus single-probands using the two-tailed Wilcoxson matched-pairs signed rank test.

By default, GEM evaluates every gene and transcript for the presence of overlapping SVs. Notably, four benchmark cases did not include externally called SVs in their input VCFs (these had been previously diagnosed by manual inspection and orthogonal confirmatory tests; **Table 2**). Nevertheless, GEM inferred the existence of these four SVs using its *ab initio* SV identification algorithm, and evaluated them jointly with SNVs and indels (“Methods”). To further demonstrate this innovative functionality, we removed all external SV calls from each input VCF file of the 14 WGS cases (as GEMs *ab initio* SV imputation is currently limited to WGS data), and reran GEM. GEM re-identified 13 of the 14 of the causative SVs. Athough GEM’s inferred SV termini were imprecise, an overlapping SV of the same class (duplication, deletion, or CNV) and ploidy to that in the original VCF was inferred, and the same high scoring gene and mode of inheritance/genotype (autosomal dominant, simple recessive, or compound heterozygote) was ranked 1^st^. SV recall within the top 1, 5, and 10 ranked GEM results were 71%, 86%, and 93%, respectively. The single false negative was a small (4kb) homozygous deletion. GEM failed to identify this SV because it did not span sites with known variation in the gnomAD database [46], upon which *ab initio* SV inference is based (“Methods”). With regard to specificity, the mean and median number of results with genes with BF > 0 in these cases was 10.6 and 12.5, respectively. These values differed only slightly from the results obtained using external SV calls (8.7 and 9.5, respectively), despite the fact every gene and transcript was evaluated for the presence of SVs.

Collectively, these results demonstrate the accuracy of GEM’s *ab initi*o approach to identification and prioritization of SVs without recourse to external calls and databases of known causative SVs. Thus, GEM compensates, in part, for the low recall of SVs from short-read sequences. If an external SV calling pipeline fails to detect an SV, there is still the possibility that GEM will identify it via this *ab initio* approach. This capability, together with GEM’s ability to accurately prioritize SVs in the context of SNVs and short indels, addresses an unmet need for clinical applications. This characteristic also makes GEM well suited for re-analyses older cases and/or pipelines lacking SV calling.

### Leveraging automated phenotyping from clinical natural language processing

Ontology-based phenotype descriptions, using Human Phenotype Ontology (HPO) terms [43], are widely used to communicate the observed clinical features of disease in a machine-readable format. These lists of terms are usually derived by manual review of patient EHR data by trained personnel, a time-consuming, subjective process. A solution is automatic extraction of patient phenotypes from clinical notes using clinical natural language processing (CNLP) [28,56]. One challenge has been that CNLP generates many more terms than by manual extraction. Thus, manual curation yielded an average of 4 HPO terms (min = 1, max = 12) in the benchmark cohort, while CNLP yielded an average of 177 HPO terms (min = 2, max = 684). Some of the extra CNLP terms are hierarchical parent terms of those observed, raising the concern that their inclusion diminishes the average information content in a manner that could impede diagnosis [27]. To investigate the effect of CNLP-derived HPO terms on GEM’s performance, we analyzed the benchmark cohort both with HPO terms extracted by commercial CNLP (“Methods”) and manually extracted HPO terms.

**Fig. 2** shows the distributions and medians for ranks and GEM gene scores of true positives, as well as the number of gene candidates with BF ≥ 0.69 (moderate support), for manual and CNLP terms. The median rank of the causal gene with GEM did not significantly differ with CNLP- and manually-derived phenotype descriptions (**Fig. 2A**). The median GEM gene score with CNLP-derived phenotypes of true positives was higher than with manual phenotypes (**Fig. 2B**). The number of candidates above the BF threshold was higher with manual phenotypes than CNLP (**Fig. 2C**). CNLP rescued a few true positives with low ranks and negative BF scores compared to manual phenotype descriptions (**Fig. 2A** and **B)**. These results demonstrate that GEM performs somewhat better with CNLP-derived phenotype descriptions as part of an automated interpretation workflow, than with sparse, manual phenotypes.

### Resilience to mis-phenotyping and gaps in clinical knowledge

Given the potentially noisy nature of the CNLP phenotype descriptions, it was important to examine the sensitivity of GEM is to mis-phenotyping. To address this question, we randomly permuted CNLP-extracted HPO terms between cases, weighting by term frequency within the cohort, so that every case maintained the same number of HPO terms as CNLP originally provided. Permuting HPO terms resulted in lower gene scores, and several cases would have been lost for review had the gene score threshold of BF ≥ 0 still been used, but ranks are unaffected (98% in top 10**; Fig. 3**). This represented lower bound estimates, as actual mis-phenotyping (short of data tracking issues) would be much less. It is also worth noting that even using randomly permuted phenotype descriptions, GEM’s performance still exceeded that of Phevor and Exomiser using the correct phenotypes (**Supp. Fig. 2**). We therefore conclude that GEM is resilient to mis-phenotyping.

We also evaluated the impact of gaps in clinical knowledge on GEM performance by withdrawing annotations from a key clinical database, ClinVar. Absence of ClinVar annotations had minimal impact in ranking, although it reduced median gene scores (1.1 vs. 2.7), resulting in 9 cases no longer meet the minimum Bayes Factor threshold ≥ 0 (any support; **Fig. 3**). Clearly, ClinVar provided GEM with valuable information. Nonetheless, without ClinVar, GEM’s top 10 maximal recall (88%) still exceeded that of Phevor (72%) and Exomiser (65%; **Fig. 1**). More broadly, these results show that integrating more datatypes in GEM improves diagnostic performance and results in greater algorithmic stability (**Figs. 2** and **3**).

### GEM performs equivalently on parent-offspring trios and single probands

Parent-offspring trios are widely used for molecular diagnosis of rare genetic disease. While a recent study showed that singleton proband sequencing returned a similar diagnostic yield as trios [57], interpretation of trio sequences is less labor intensive. For example, trios enable facile identification of *de novo* variants, which is the leading mechanism of genetic disease in outbred populations [58]. Likewise, in recessive disorders, proband compound heterozygosity can be automatically distinguished from two variants in *cis*. However, these benefits are associated with increased sequencing costs. Moreover, both parents are not always available for sequencing or do not wish to have their genomes sequenced.

To understand how GEM performs in the absence of parental data, we reanalyzed the 63 trio and duo cases from the benchmark cohort as singleton proband cases. Surprisingly, we observed insignificant differences in the mean rank of the causal gene (**Fig. 4A**), GEM score of the causal gene (**Fig. 4B**), or number of candidates with BF ≥ 0.69 (**Fig. 4C**), using either using manually or CNLP extracted HPO terms. In contrast, this re-analysis was associated with a decline in the performance of Exomiser (**Supp. Fig. 6)**. These analyses demonstrated that GEM was resilient to the absence of parental genotypes, a feature that could increase the cost effectiveness and adoption of WGS.

**Figure 6.**
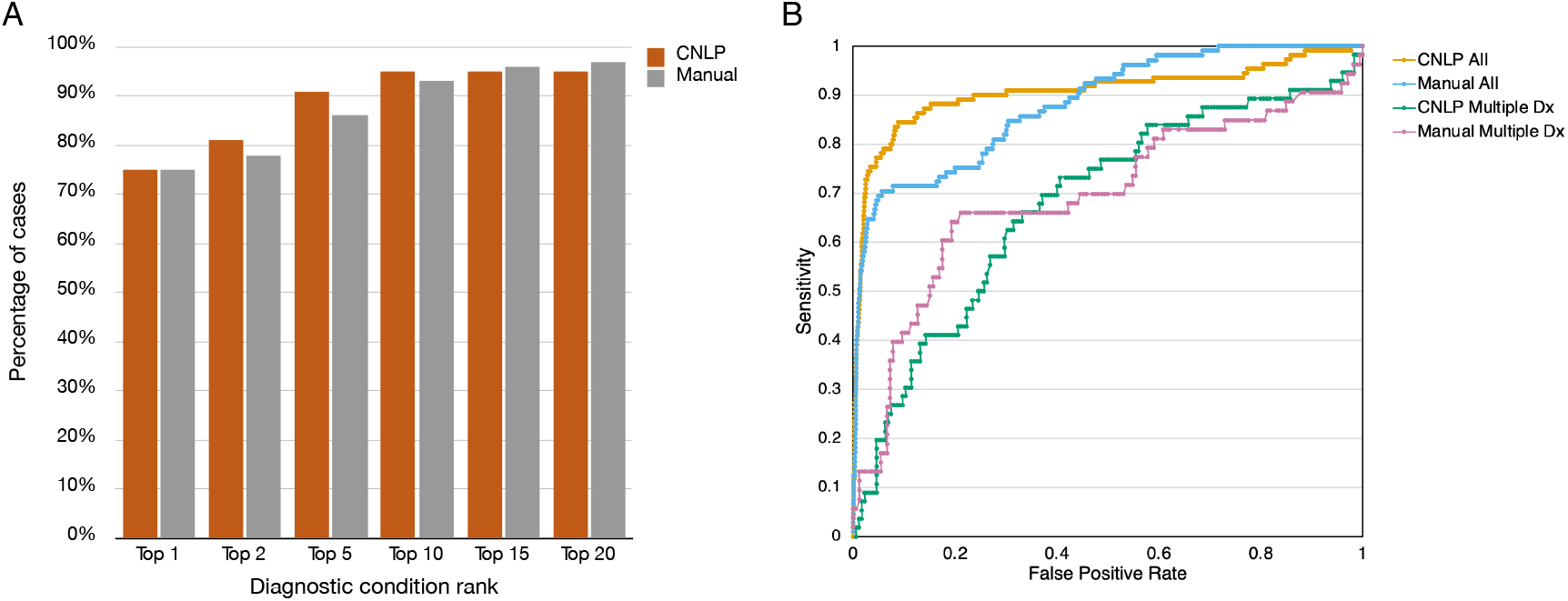
Performance of GEM condition-match scores for diagnostic nomination. (**A**) Ranks for reported diagnostic conditions for the benchmark dataset, using a GEM gene BF score ≥ 0.69 and sorted by CM score, for HPO terms derived from CNLP or manual curation. (**B**) Receiver-operator characteristic curves for the condition-match (CM) score for all hits with BF *≥* 0. CNLP All: HPO extracted from clinical notes by CNLP; AUC=0.91. Manual: Manually curated HPO terms; AUC=0.88. CNLP Multiple Dx: CNLP-derived CM score for the true positive disorder versus the other possible disorders associated with that gene; AUC=0.68. Manual Multiple Dx: As for CNLP-derived CM, but using manually curated HPO terms; AUC=0.69.

### GEM scores optimize case review workflows

Conventional prioritization algorithms rank variants, enabling manual reviewers to start with the top ranked variants, and work their way down in the list until a convincing variant is identified for further curation, classification, and possible clinical reporting. This review process typically involves: a) assessing variant quality, deleteriousness, and prior clinical annotations; b) evaluating whether there is a reasonable match between the phenotypes exhibited by the patient and those reported for condition(s) known to be associated with defects in the corresponding gene, and c) considering the match in mode(s) of inheritance reported in the literature for the candidate disease and the patient’s diplotype.

GEM accelerates this process, because it intrinsically considers variant quality, deleteriousness, prior clinical annotations, and mode of inheritance. Furthermore, at manual review, GEM gene scores summarize the relative strength of evidence supporting the hypothesis that the gene is damaged and that this explains the proband’s phenotype. GEM scores provide a logical framework for setting thresholds with regard to the optimal number of candidates that should be reviewed to achieve a desired diagnostic rate. This enables laboratory directors and clinicians to dynamically set optimal tradeoffs of interpretation time and diagnosis rate for specific patients, relative to their suspicion of a genetic etiology or results of other diagnostic tests.

We examined the effect of different BF thresholds on recall (true positive rate) and median number of gene candidates for review in the benchmark cohort (**Fig. 5)**. In such analyses, it is germane to consider the concept of *maximal true positive rate (or recall)* to measure the theoretical proportion of true positive diagnoses recoverable by perfect interpretation when reviewing a set of *N* genes containing the true positive. For example, in the benchmark dataset, a GEM causal gene score threshold ≥ 0 would retain a median of ten candidates for review and provide a 99% maximal recall; whereas a threshold of ≥ 0.5 would retain a median of four candidates for review for a 97% maximal recall (**Fig. 5**).

These results illustrate how a tiered approach to case review using GEM gene scores could minimize the number of candidate genes to review, and, thereby manual interpretation effort. For example, a first pass review of candidates with a gene BF ≥ 0.69 provided an expected 95% diagnostic rate (and a corresponding median of 3 genes to be manually reviewed). If followed by a second pass using a threshold > 0, if no convincing candidates are found, an additional 4% possible diagnoses would be recovered, involving review of a median increment of seven genes. Application of this two-tiered approach to the benchmark dataset of 119 cases (**Fig. 1**), required manual final review of 395 candidate genes (3 genes in 115 cases and 10 genes in 5 cases), or an average of 3.3 candidate genes per case, for a maximal recall of 99%. Finally, review of candidates with BF <0 recovered the last true positive in the benchmark cohort (*COL4A4*, ranked 40^th^ in the GEM report with a BF=-0.6. This case was a phenotypically and genotypically atypical autosomal dominant presentation of Alport syndrome 2 (MIM 203780).

### Clinical Decision Support for Diagnosis

Quantifying how well the observed phenotypes in a patient match the expected phenotypes of Mendelian conditions associated with a candidate gene is challenging for clinical reviewers and is a major interpretation bottleneck. In practice, clinicians look for patterns of phenotypes, biasing their observations. In addition, patient phenotypes evolve as disease progresses. And there is considerable, disease-specific heterogeneity in the range of expected phenotypes. Simply comparing exact matches of the patient’s observed HPO terms with those expected for that disease is suboptimal, because the observed and expected HPO terms are often hierarchical neighbors, rather than exact matches. Missing terms, particularly those considered pathognomonic for a condition, and “contradictory” terms further complicate such comparisons by clinicians. Thus, generation of quantitative, standardized, unbiased models of disease similarity has proven elusive. GEM can automate or provide clinical decision support for this process via a condition-match (CM) score (“Methods”). The GEM CM score summarizes the match between observed and expected HPO phenotypes for genetic diseases, and considers the known mode(s) of inheritance, associated gene(s), their genome location(s), proband sex, the pathogenicity of observed diplotypes, and ClinVar annotations. Importantly, CM scores reflect relationships between phenotype terms as expressed in the HPO ontology graph, enabling inclusion of imprecise matches in similarity comparisons. CM scores can be used in a wide variety of clinical settings to prioritize and quickly assess possible Mendelian conditions as candidate diagnoses, a process we term *diagnostic nomination*.

Specific, definitive, genetic disease diagnosis remains a significant challenge for clinical reviewers, even with the short, highly informative candidate gene lists provided by tools such as GEM. This is because many genes are associated with more than one Mendelian disease. For example, application of a GEM causal gene score threshold *≥* 0.69 to the 119 benchmarking probands results in a median of 3 gene candidates per proband (c.f. **Fig. 5**), associated with a maximal gene recall of 95%. However, because many genes are associated with more than one disease, clinical reviewers would actually need to consider around 12 candidate Mendelian conditions per proband (data not shown). This difficulty is exacerbated by the fact that most laboratory directors are not physicians and lack formal training in clinical diagnosis.

Determination of a specific, definitive genetic disease diagnosis among several candidates can be accomplished with a combination of GEM CM scores and causal gene scores (**Fig. 6**). Using the benchmark cohort’s true (reported) gene and disorder diagnoses as ground truth, we used a GEM gene score threshold ≥ 0.69 to recover gene candidates, and the associated CM scores to rank order the diseases associated with those gene candidates (**Fig. 6A)**. Using CNLP-derived phenotypes, the true disease diagnosis was the top nomination by CM score in 75% of cases, within the top 5 in 91% of cases, and within the top 10 in 95% of cases. Performance was inferior with manually extracted phenotype terms. The area under the receiver-operator characteristic (ROC) curves (AUCs) were 0.90 and 0.88, for CNLP and Manual terms, respectively (**Fig. 6B)**. This implied that the larger number of CNLP extracted terms conveyed greater information content, permitting better discrimination of the correct diagnostic condition, than sparse, manually extracted phenotypes [27].

In the benchmark dataset, 58 of the 100 candidate genes (excluding cases with causal, multigenic SVs) were associated with 2 or more disorders (median of 3 gene-disorder, maximum of 15; **Supp. Fig. 7** shows the example of *ERCC6*). We measured how well the CM score distinguished between multiple, alternative disorders associated with the same gene (**Fig. 6B**). In these 58 cases, the AUC was less than that for CNLP with the entire set of candidate genes in the benchmark dataset (0.68 vs 0.9). This decrease can be at least partially explained by the high similarity (and in some cases identity) of the clinical features of different disorders associated with the same gene. Thus, a combination of GEM gene and CM scores can refine candidate disorders for clinical reporting, further reducing review times.

### Re-analysis of previously undiagnosed cases

Recent reports show that re-analysis of older undiagnosed cases suspected of rare genetic disease can yield new diagnoses supported by incremental increases in knowledge of pathogenic variants, disease-gene discoveries, and reports of phenotype expansion for known disorders [59,60]. While worthwhile, there are barriers to re-analysis, such as limited reimbursement and low incremental diagnostic yield, that limit use to physician requests. Ideally, all negative cases would be re-analyzed automatically periodically, and a subset with high likelihood new findings would be prioritized for manual review. The strong correlation between true positive rates and GEM gene scores (**Fig. 5**) suggested a strategy for triaging re-analyzed cases for manual review: only cases for which the recalculated GEM score had increased sufficiently to suggest a high probability of a new diagnosis would pass the threshold for manual review. Likewise, GEM condition match scores could be used to search all prior cases to identify the subset of undiagnosed cases with support for particular Mendelian conditions, aiding cohort assembly for targeted re-analysis based upon particular proband phenotypes, or for review by particular medical specialists. Of note, an advantage of CNLP is that it is possible to automatically generate a new clinical feature list at time of re-analysis. This is particularly important in disorders whose clinical features evolve with time and where the observed features may be nondescript at presentation.

To test the utility of GEM for re-analysis, we selected 14 difficult cases that had received negative WGS by RCIGM. For these reanalyzes, we used CNLP-derived HPO terms (**Table 3**) and a more stringent gene BF threshold ≥ 1.5 to restrict the search to very strongly supported candidates. Ten cases yielded no hits. Four cases returned a total of 7 candidate genes. Review of three cases did not return new diagnoses. In the remaining case, a new likely diagnosis was made of autosomal dominant Shwachman-Diamond Syndrome (MIM: 260400) or severe congenital neutropenia (MIM: 618752) [61,62], both of which are associated with pathogenic variants in *SRP54*. The respective CM scores using 261 CNLP-derived terms were relatively high (0.893 and 0.672, respectively). The association of *SRP54* and these disorders was first reported in November 2017 [61] and entered in OMIM in January 2020 [63], which explained why it was not identified as the diagnosis originally in July 2017. Assuming *de novo* occurrence, the identified p.Gly108Glu variant was classified as likely pathogenic (meeting PM2, PM1, PP3 and PM6 of the ACMG guidelines). This was a singleton proband sequence and confirmation is being pursued. Thus, GEM re-analysis of 14 negative cases led to 7 gene-disorder reviews (an average of 0.5 per case), and yielded one likely new diagnosis, which was consistent with prior re-analysis yields [59,60].

**Table 3.** Previously undiagnosed cases with potential leads at GEM gene score BF > 1.5 out of 14. Zygo – zygosity; Hom – homozygous; Het -heterozygous; Dup – large duplication.

| Case | Pedigree | Sex | Assay | Rank | Chr | Gene | Variant Type | Variant ACMG | *de novo* | Zygo | GEM score | Mode of Inheritance | MIM ID(s) | CM Score(s) |
| --- | --- | --- | --- | --- | --- | --- | --- | --- | --- | --- | --- | --- | --- | --- |
| UD1 | Single | Male | WGS | 1 | 14 | SRP54 | SNV | Likely Pathogenic | Likely | Het | 1.76 | Autosomal dominant | 618752, 260400 | 0.672, 0.893 |
| UD2 | Trio | Male | WGS | 2 | X | GK | SNV | VUS | Yes | Het | 1.60 | X-linked recessive | 307030 | 1.119 |
| UD2 | Trio | Male | WGS | 3 | 16 | FANCA | SNV | VUS | No | Hom | 1.55 | Autosomal recessive | 227650 | 1.315 |
| UD3 | Single | Male | WGS | 1 | 16 | TSC2 | Dup | VUS | Likely | Het | 1.64 | Autosomal dominant | N/A | N/A |
| UD4 | Single | Male | WGS | 1 | 1 | SLC25A24 | SNV | VUS | Likely | Het | 1.86 | Autosomal dominant | 612289 | 1.995 |
| UD5 | Trio | Female | WGS | 1 | X | STAG2 | SNV | Likely Pathogenic | Yes | Het | 1.53 | X-linked dominant | 301022 | 1.25 |

## Conclusions

Here we described and benchmarked a Bayesian, AI-based gene prioritization tool for scalable diagnosis of rare genetic diseases by CNLP and WES or WGS. [31][19,27,28,64,65]GEM improved upon prior, similar tools [19,27,28,64,65] by incorporating OMIM, HPO and ClinVar knowledge explicitly, automatically controlling for confounding factors, such as sex and ancestry, compatibility with CNLP-derived phenotypes, SVs and singleton probands, and by directly nominating diplotypes and disorders, rather than variants.[19,27,28,64,65]

In the cohorts examined, GEM had maximal recall of 99%, requiring review of an average of 3 candidate genes, and less than one half of the associated disorders nominated by other widely used variant prioritization methods per case. Improved diagnostic performance is anticipated to enable faster and more cost-effective, tiered reviews. These features hold promise for reduced time-to-diagnosis and greater scalability for critical applications, such as in seriously ill children in the NICU/PICU [27,66].

Uniquely, GEM provided AI-based unified gene prioritization for SVs and small variants. Hitherto, this has been frustrated by the high false positive rates of SV calls in short read sequences and lack of a suitable framework for AI-based SV pathogenicity assertions [53,54]. Furthermore, GEM inferred SV calls *ab initio* from WGS when they were not provided. These functionalities are critical for re-analyzing older cases, and for pipelines lacking SV calls.

Finally, in a small data set, we showed that GEM can efficiently re-analyze cases, potentially permitting cost-effective, scalable re-analysis of previously negative patients as disease, gene and variant knowledge evolves [60,67]. Indeed, integration of GEM and CNLP could enable automatic surveillance for rare disease patients [68] from genomes obtained for research or other clinical tests performed in healthcare [69,70].

## Data Availability

The datasets supporting the conclusions of this article are included within the article and its additional files. Due to patient privacy, data sharing consent, and HIPAA regulations, our raw data cannot be submitted to publicly available databases. GEM, PHEVOR and VAAST software implementations for versions used in this analysis are part of the Fabric Enterprise analysis platform and is commercially available (https://www.fabricgenomics.com). Exomiser source code (version 12.1.0) is available on GitHub at: https://github.com/exomiser/Exomiser.

## Availability of data and materials

The datasets supporting the conclusions of this article are included within the article and its additional files. Due to patient privacy, data sharing consent, and HIPAA regulations, our raw data cannot be submitted to publicly available databases. Anonymized outputs from GEM, Phevor, VAAST, and Exomiser for the benchmark dataset cases are tabulated in Additional file A, and GEM for the validation dataset cases in Additional file B. Condition match scores for hits with gene BF >0 used for Figure 6 are tabulated in Additional file C. GEM, PHEVOR and VAAST software implementations for versions used in this analysis are part of the Fabric Enterprise analysis platform and is commercially available (https://www.fabricgenomics.com). Exomiser source code (version 12.1.0) is available on GitHub at: https://github.com/exomiser/Exomiser.

## Acknowledgements

We thank Jeff Rule and Birgit Crain for help in extracting case data for the RCH cases from Fabric Enterprise, Brent Lutz for project management, and Sandy White for interinstitutional coordination (Fabric Genomics, Oakland CA). We are grateful to Joe Azure, Josh Grigonis, Jeff Rule, Bjoern Achilles-Stade, Peter Spiro (Fabric Genomics, Oakland, CA), Ray Drummond, Richard Littin, Aidan Scarlet, Mike Richdale, and Tim Dawson (NetValue Ltd, Hamilton, New Zealand) for software and architecture development efforts to implement GEM in the Fabric Enterprise platform. We also acknowledge Edward Kiruluta and Marco Falcioni (formerly Fabric Genomics) for useful discussions early on the project.

## Author’s Contributions

FV, MY, and SC designed the study and analysis strategy. MY developed the algorithms. FV provided algorithm requirements, designed UIs, and led the software implementation. BM and EF implemented analysis pipelines. BM, EF, EJH, JM, FV and MY performed data analysis. AF, AHB, BB, BSL, CAB, CAG, JM, KJ, KÕ, KR, LG, MH, MN NV, PBA, SC, SP, TW, and VN complied cases and clinical evidence. PB provided feedback on features and development. MGR and SK sponsored the project and provided helpful discussions and edits of the manuscript. FV and MY wrote the manuscript. All authors reviewed and suggested edits for the final version of the manuscript.

## Supplementary Figures

**Supplemental Figure 1.**
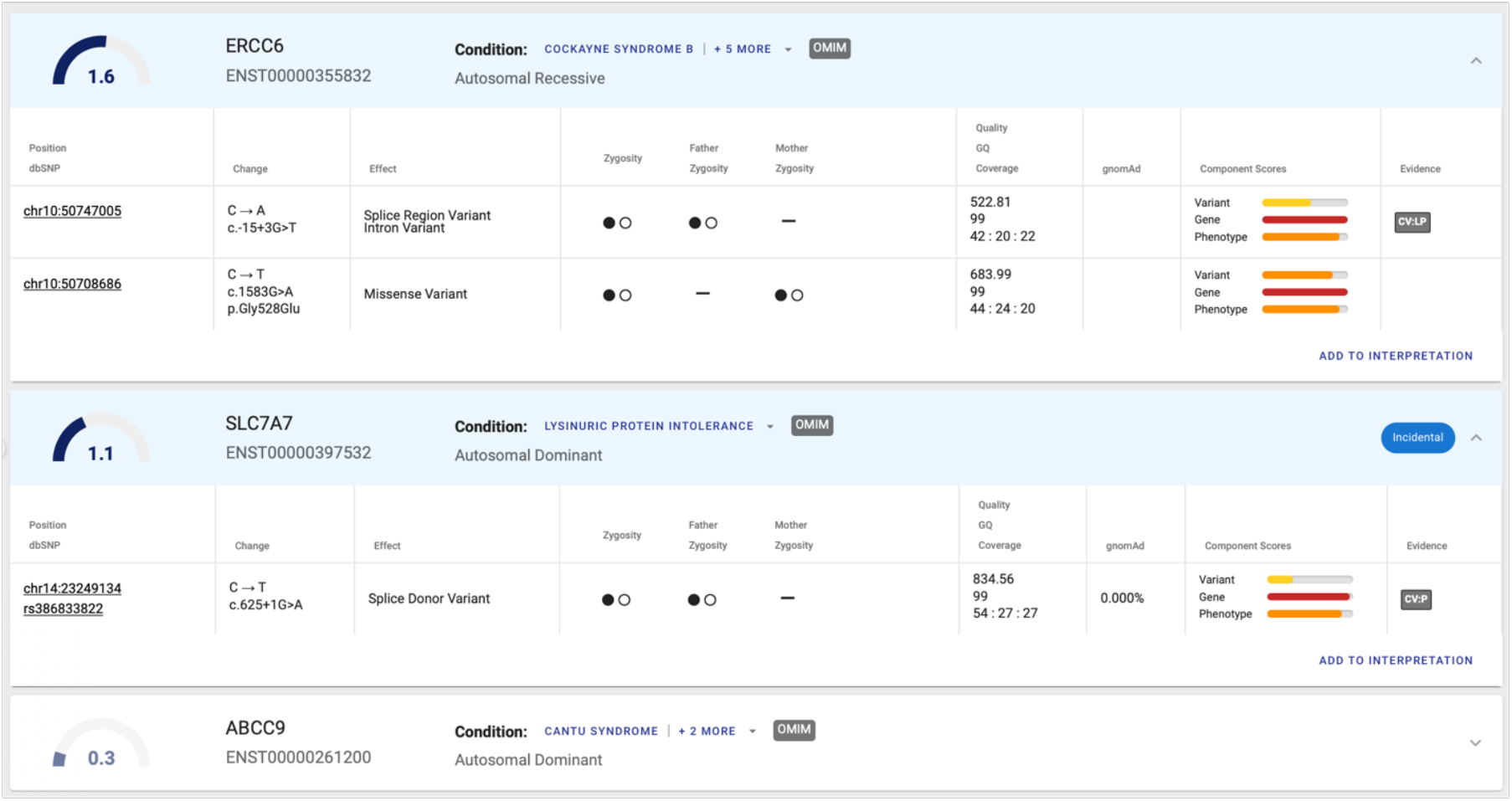
Example of GEM output. Each card represents a gene scored by GEM, sorted by the Bayes Factor gene score (large number top left and ideogram), diplotypes are automatically grouped together in a card, and their composite score is shown in the upper left-hand corner of the card. GEM outputs also include several key parameters that provide additional guidance for case review, improving explainability and speeding interpretation, including: gene symbol; canonical transcript ID; mode of inheritance selected by GEM; disease conditions associated with gene (drop-down menu); position of variant; sequence change and HGVS notation; effect of variants in transcript; genotype zygosity in proband and parents (if available); variant quality (QUAL), GQ, coverage, and allele counts; gnomAD allele frequencies (if available); major components scores feeding into GEM: a) ‘Variant’, variant deleteriousness score (VVP), b) ‘Gene’, gene burden score (renormalized VAAST score), and c) ‘Phenotype’, gene phenotype prior (renormalized Phevor gene priors) – all scaled 0-1 and presented logarithmically as color coded bar for ease of presentation; and ‘Evidence’, ClinVar pathogenic/likely pathogenic evidence, if available. A badge labels a card “incidental,” if, although score is high, mode of inheritance may not match what is known about the associated conditions, i.e. the individual is a carrier of a known pathogenic variant.

**Supplemental Figure 2.**
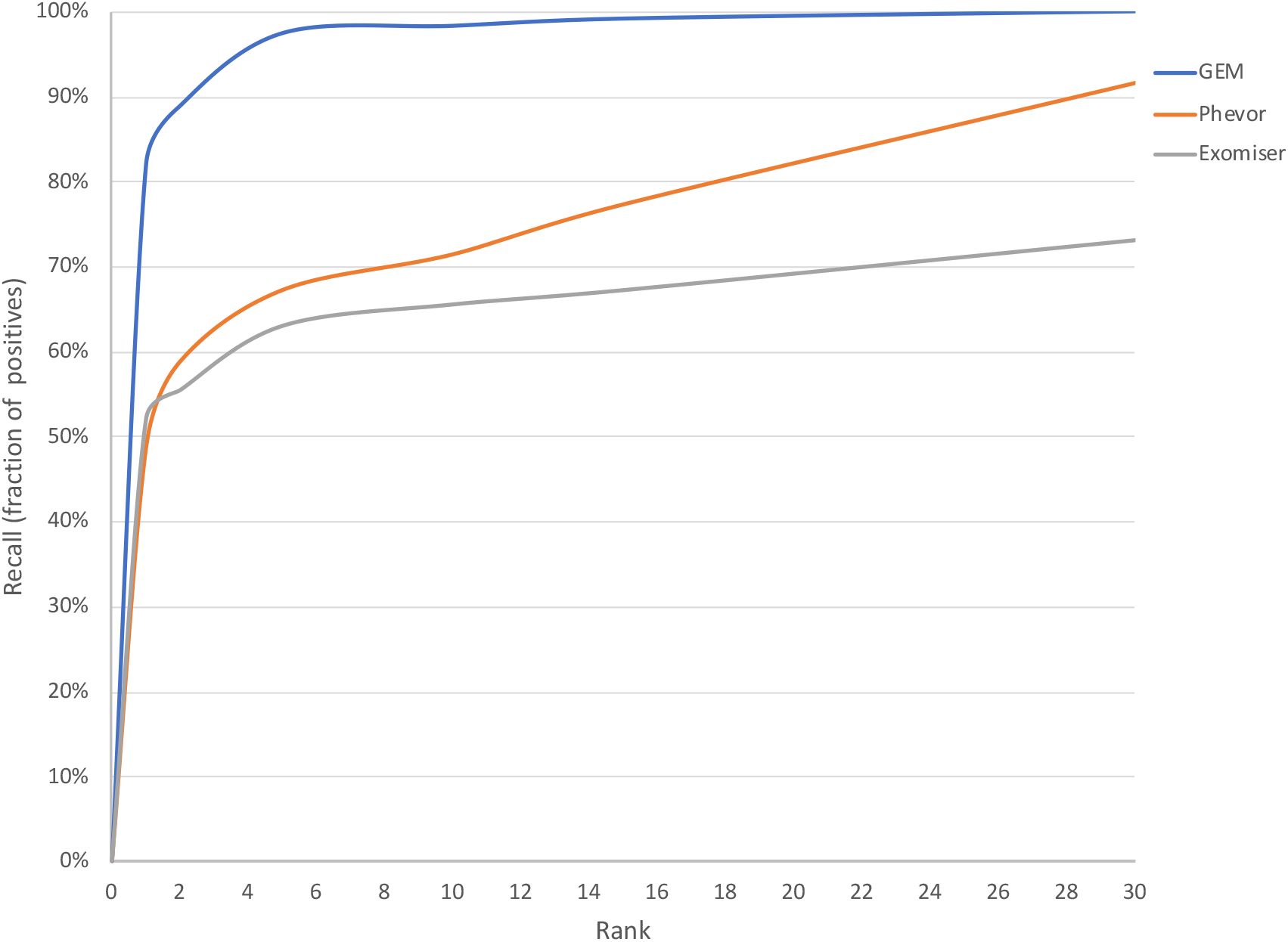
Cumulative ranking of benchmark cases by GEM, Phevor, and Exomiser. Percentage of true causal gene (or variant in the case of causal SVs) identified across each rank in the output list the entire benchmark cohort of 119 cases of the benchmark cohort. Patient phenotypes curated by clinicians manually from medical records expressed as HPO terms where provided as inputs to GEM, Exomiser and Phevor. It should be noted that GEM ranks correspond to genes, which may include one or two variants (the latter in the case of a compound heterozygote), whereas the ranks for the other methods are for variants; in the case of compound heterozygotes we indicate the rank of the top-ranking variant for these methods. Ranks were truncated at 30, although Phevor’s last rank was 356 whereas Exomiser was 112. Note that Exomiser did not provide true positives in its output for 18 cases and VAAST for 4 cases.

**Supplemental Figure 3.**
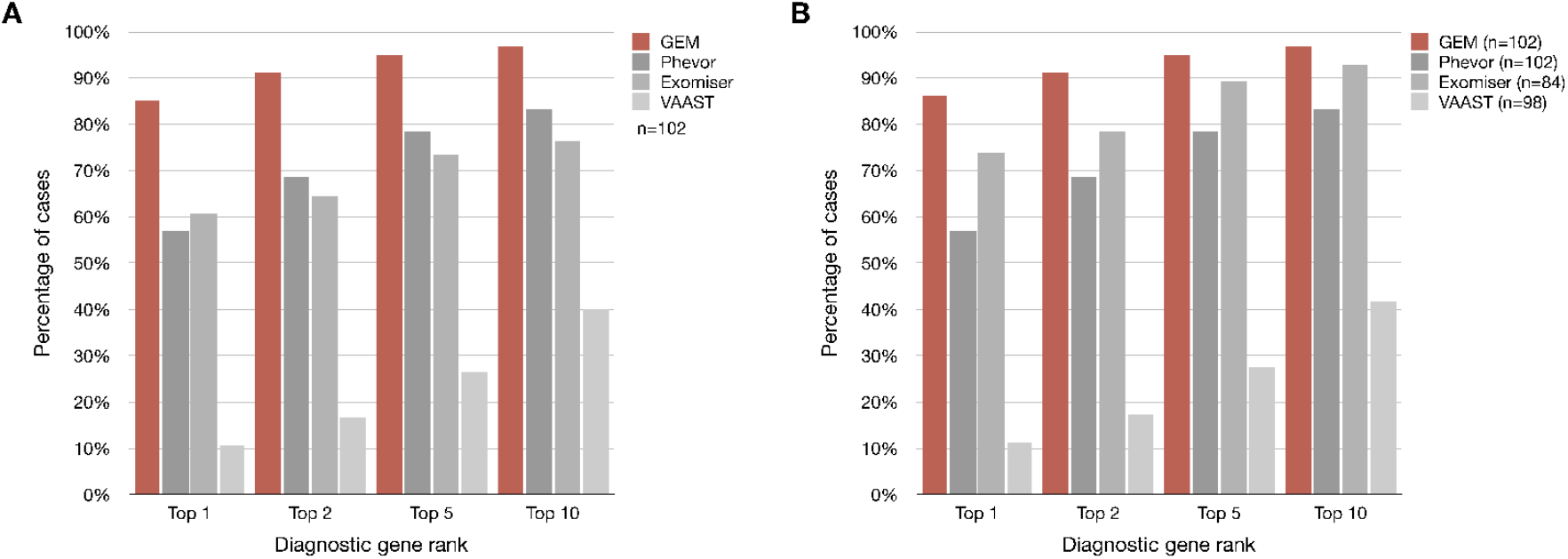
GEM AI-CDS outperforms variant prioritization approaches. Comparative performance of GEM’s ability to rank diagnostic genes versus those derived from previous variant prioritization algorithms. (A) Percentage of the 102 cases from the benchmark cohort, where SVs were not causal, is identified within the top 1st, 2nd, 5th, or 10th genes by the different methods compared: GEM, Phevor, Exomiser, and VAAST. For the former three methods, patient phenotypes curated by clinicians manually from medical records expressed as HPO terms where provided as inputs; VAAST only considers variant information. It should be noted that GEM ranks correspond to genes, which may include one or two variants (the latter in the case of a compound heterozygote), whereas the ranks for the other methods are for variants; in the case of compound heterozygotes we indicate the rank of the top ranking variant for these methods. (B) Since Exomiser and VAAST were unable to rank the causal variant in a number of cases, here the comparison uses as denominator only the cases each method was able to provide a rank for a causal gene/variant; this should be considered the best possible scenario for each method.

**Supplemental Figure 4.**
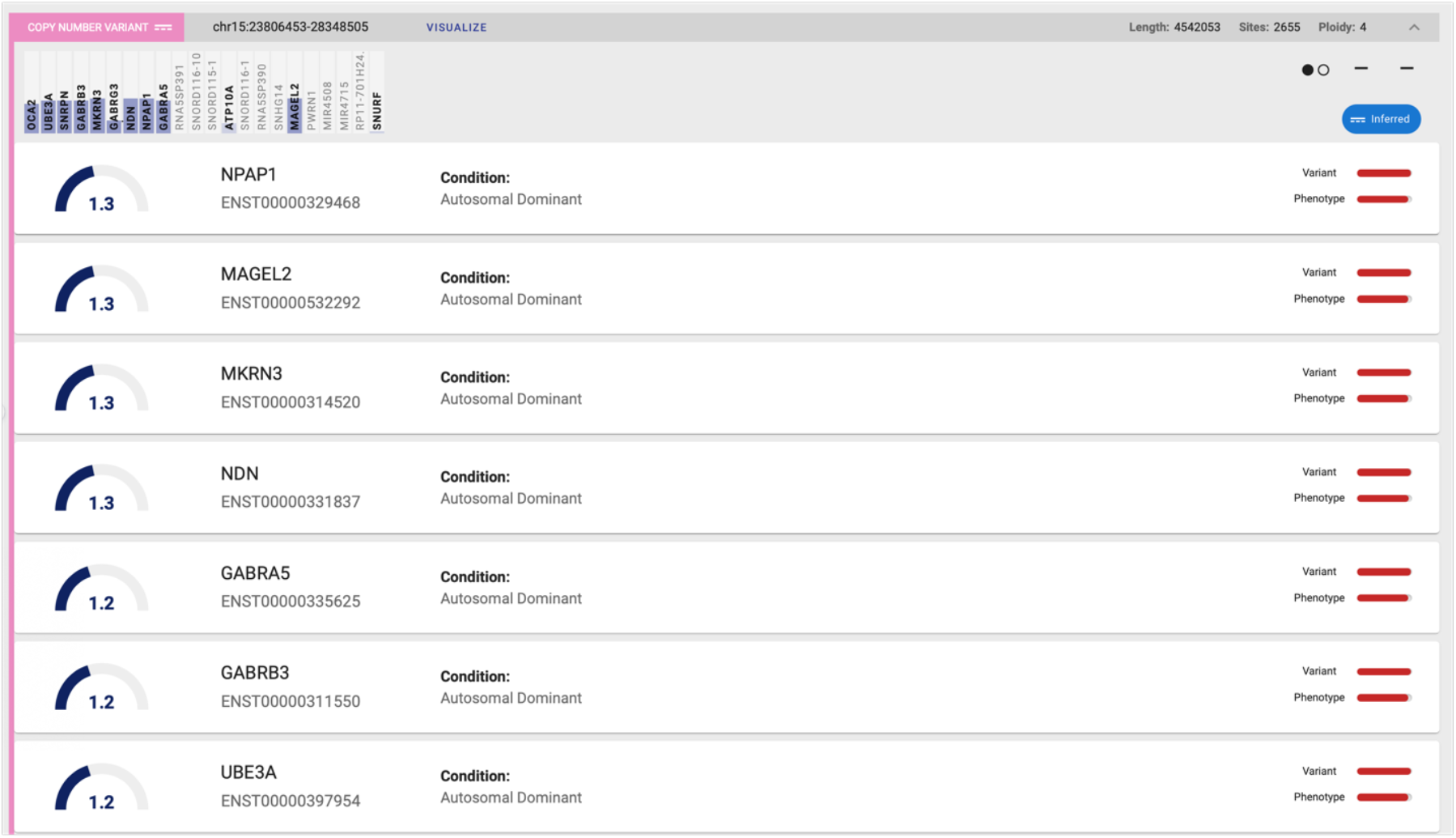
GEM report with scored SV example. GEM cards can also represent structural variants and contain overlapped genes scored by GEM and meeting a minimum bayes factor score. Genes within the SV are ranked, and the card can be expanded to visualize an individual gene harbored (as shown) and is listed in the rank of the gene with the highest bayes factor score (“anchor” gene). GEM specifies the type of event (deletion, duplication, copy number change for ploidy greater than 3; genomic coordinates; an ideogram showing the genes overlapped by the event and those genes that were scored by GEM (the height of the bar a succinct representation of the gene Bayes Factor); the length of the event in base pairs; number of SNVs with gnomAD allele frequency data supporting the event (whether the variants were provided on the input VCF or internally inferred ab initio by GEM, like in this case); and estimated ploidy from the data. Component scores shown include a variant deleteriousness score, and the Phevor gene prior, but not burden score, as VAAST cannot presently calculate burden for SVs. The card also indicates if the SV was internally inferred (“Inferred” badge above right). If GEM scores an SV in compound heterozygote state with an SNV or small indel, then the gene card will show details of the small variant as usual. Links are provided for further visualization of the event in the context of annotations relevant to structural variation interpretation such as Decipher Syndromes and ISCA microdeletion/duplication database.

**Supplemental Figure 5.**
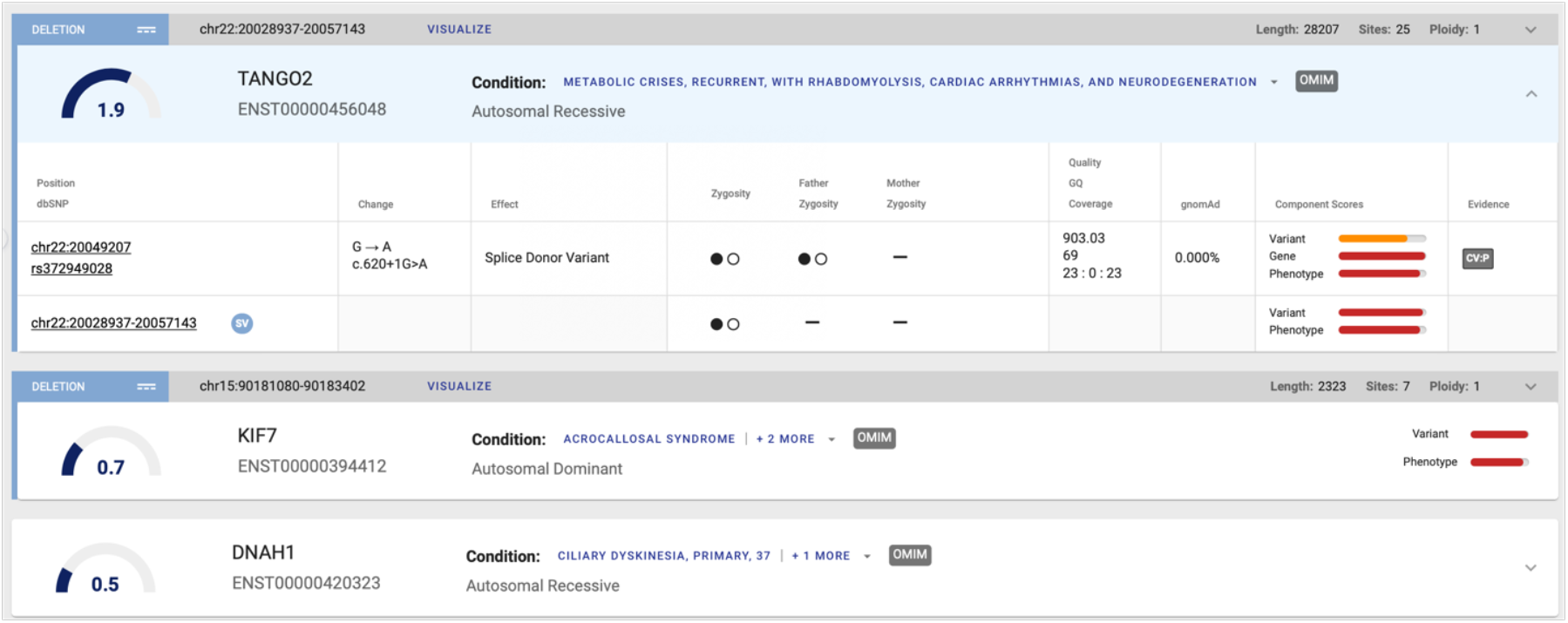
Example of compound heterozygote genotype combining an SV (an ab initio inferred deletion) with an SNV overlapping the gene TANGO2. In this case the SNV shows the usual attributes, whereas the SV shows the interval of the event, its zygosity and its variant and phenotype impact scores. The GEM gene score is calculated for the compound heterozygote recessive genotype.

**Supplemental Figure 6.**
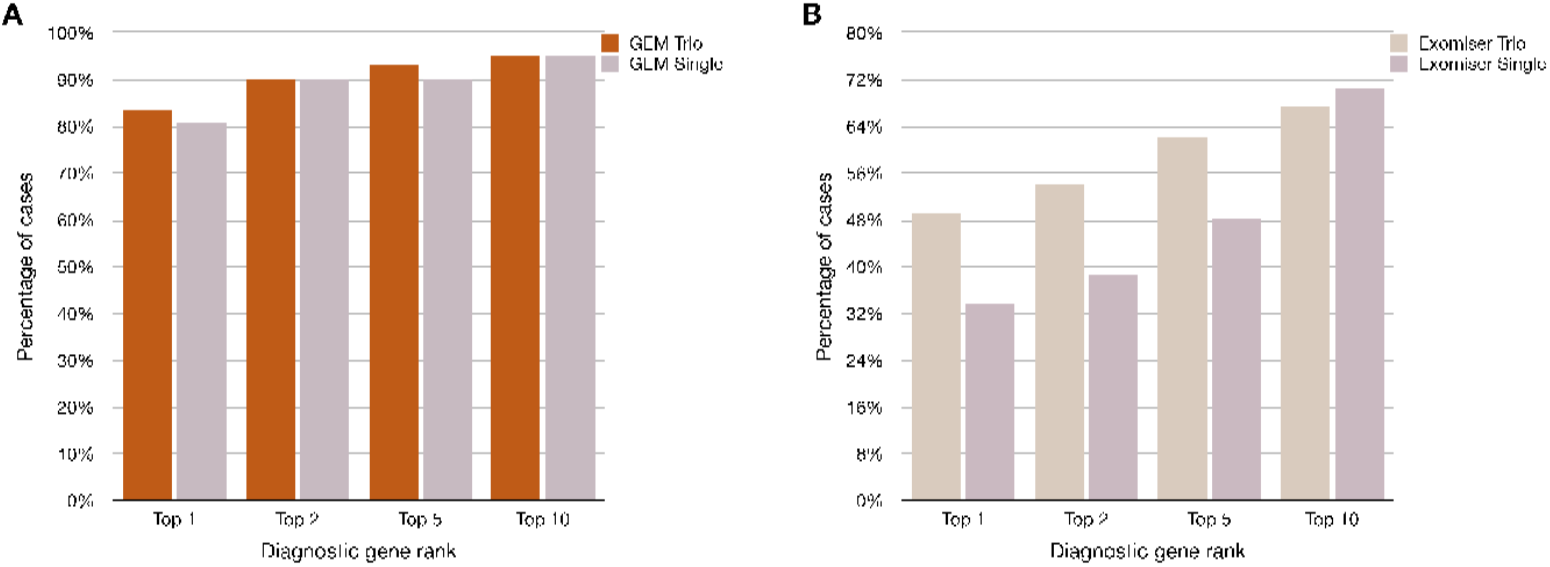
Impact of removing parent information in ranking by GEM (Panel A) and Exomiser (Panel B), showing that Exomiser ranking performance drops in the absence of parental genotypes, whereas for GEM has little impact. This analysis is restricted to 63 cases where parental information was available either as trios or duos and excluding cases with structural variation that Exomiser cannot analyze. Note difference in scale of x-axis in panels A and B.

**Supplemental Figure 7.**
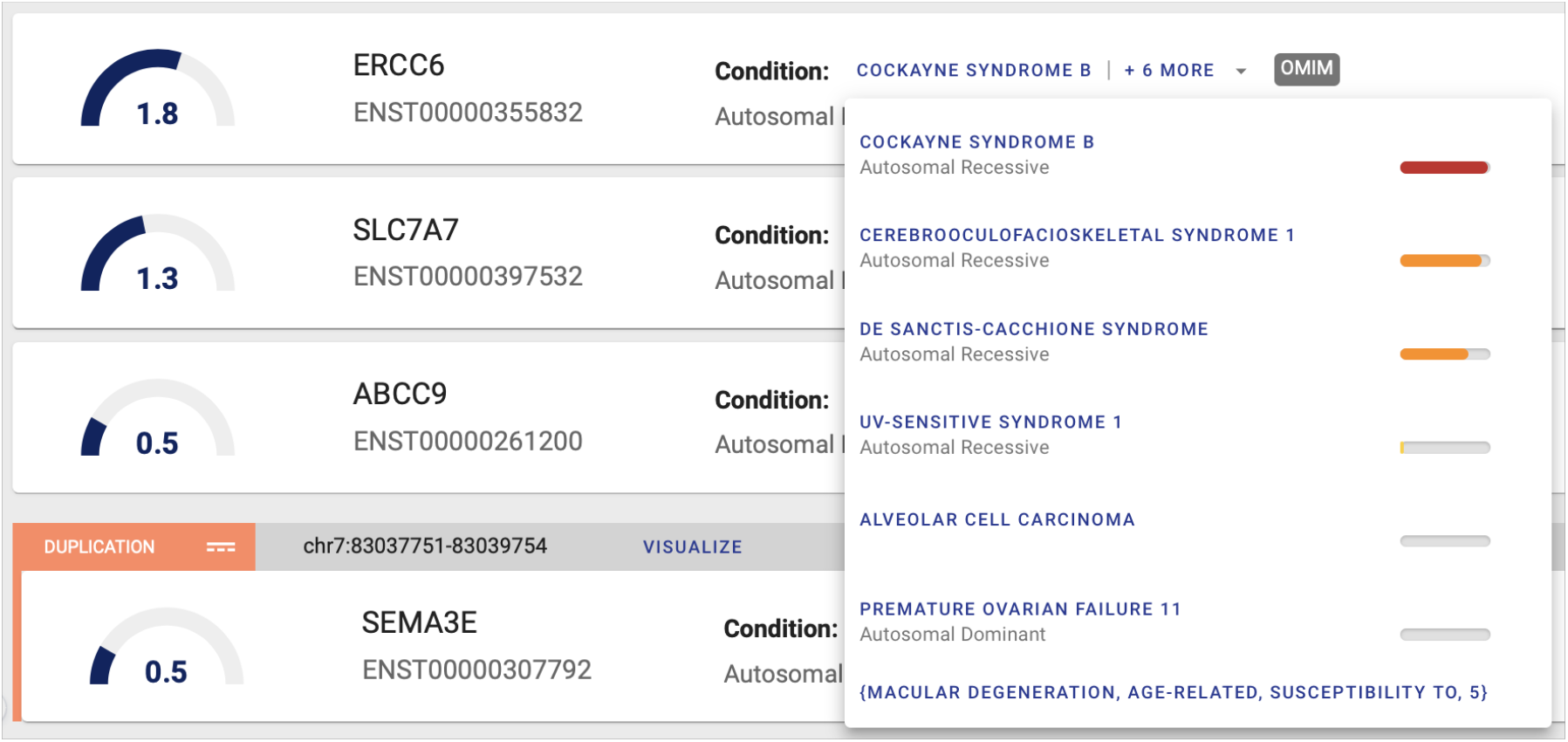
Diagnostic nomination with condition match scores in GEM report. The CM scores (color coded to represent relative magnitude and degree of Bayes Factor support) are shown next to each condition associated in a drop-down menu. The condition with the highest CM score is shown first and the remainder are sorted by the score in order of relevance as a candidate diagnosis. CM scores are normalized to 0-1 values and shown as color-coded bars next to each condition. Known mode(s) of inheritance for each condition are also noted and links to the OMIM database are provided for further information. For the case illustrated here, the gene ERCC6 was the highest ranked by GEM and is the true positive. ERCC6 is associated with 6 conditions with HPO profile and mode of inheritance, and a condition with no mode of inheritance described (not ranked by the CM score). From this display it is clear that the conditions at the bottom are poor fits to the patient phenotypes (CM scores, from the bottom: −1.42, −0.94, 0.19), whereas the top 3 can be diagnostic candidates (CM scores, from the top: 1.84, 1.46, 1.58) with GEM CM score favoring Cockayne syndrome B, the actual true positive diagnosis for this case.

